# Differential DNA Methylation of the Brain-Derived Neurotrophic Factor Gene is Observed after Pediatric Traumatic Brain Injury Compared to Orthopedic Injury

**DOI:** 10.1101/2025.06.16.25329571

**Authors:** Lacey W. Heinsberg, Aboli Kesbhat, Bailey Petersen, Lauren Kaseman, Zachary Stec, Nivinthiga Anton, Patrick M. Kochanek, Keith Owen Yeates, Daniel E. Weeks, Yvette P. Conley, Amery Treble-Barna

## Abstract

Pediatric traumatic brain injury (TBI) triggers biological changes that may differ from those observed in non-brain injuries. *BDNF* DNA methylation (DNAm) may serve as a novel, dynamic biomarker of the brain’s response and help identify TBI-specific epigenetic patterns relevant to later recovery. Therefore, the purpose of this study was to examine whether *BDNF* DNAm differed between children with TBI and those with orthopedic injury (OI, comparison group) acutely and over time. Data were derived from the Epigenetic Effects on TBI Recovery (EETR) study, a prospective, longitudinal cohort study conducted at UPMC Children’s Hospital of Pittsburgh. Children aged 3 to 18 years hospitalized at a minimum of overnight for complicated mild to severe TBI or OI without head trauma were enrolled. Exclusion criteria included prior hospitalization for TBI, pre-existing neurological or psychiatric conditions, or sensory or motor impairments precluding study participation. Blood samples were collected during hospitalization (mean=31.6 hours post-injury) and at 6 (mean=216.9 days) and 12 months (mean=405.9 days) post-injury. The primary outcome variable was DNAm assessed via pyrosequencing at five quality-controlled CpG sites in the *BDNF* gene (chromosome 11, hg38 positions 27722033, 27722036, 27722047, 27701612, and 27701614). The primary exposure was injury type (TBI vs. OI), with severity (measured via Glasgow Coma Scale [GCS]) examined as a secondary exposure within the TBI group. Primary covariates included age, sex, and race; secondary covariates included pubertal status, age-adjusted BMI, non-head injury severity, socioeconomic status, and psychosocial adversity. The final analysis sample included n=189 participants with TBI and n=105 participants with OI. Participants were 66.3% male, 83.2% White, and had a mean age of 10.6 (±4.4) years at the time of enrollment. Acutely, children with TBI showed significantly lower DNAm at three of five sites (3.17%– 5.83% lower; p=0.0044 to 6.48E-06) while controlling for age, sex, and race. One site remained significantly lower at 12 months (8.56% lower; p=0.0045); no significant differences were observed at 6 months. Observed differences remained robust across sensitivity models adjusting for secondary covariates. GCS-measured TBI severity was not associated with DNAm at any time point. These findings suggest that *BDNF* DNAm differs between children with TBI and those with OI, particularly in the acute period. *BDNF* DNAm differences may reflect early biological responses that are specific to TBI.

## 1.0 INTRODUCTION

With an estimated annual worldwide prevalence of 69 million, traumatic brain injury (TBI) is a leading cause of disability in children.^1,2^ Recovery from TBI involves a complex interplay of biological, psychosocial, and therapeutic factors. This complexity contributes to substantial heterogeneity in outcomes and remains a major barrier to developing accurate prognostic models and effective interventions.^3,4^ Although biomarker discovery has advanced the field, efforts have largely focused on protein markers of structural brain injury. While informative, these markers may not fully capture the broader biological and environmental influences that shape recovery.^5–7^ There is a critical need to identify dynamic and potentially modifiable biomarkers that reflect the child’s psychosocial environment (e.g., social drivers of health) and respond to injury progression and recovery processes over time.

Methylomic biomarkers (i.e., DNA methylation [DNAm]) are emerging as promising tools to advance our understanding of disease mechanisms, predict outcomes, and guide therapy development.^8–12^ DNAm is an epigenetic mechanism that regulates gene expression and responds dynamically to both internal (e.g., injury) and external (e.g., psychosocial adversity^13–16^, rehabilitative therapies^17–20^) factors. Preclinical^21,22^ and adult clinical^23,24^ studies show differential DNAm both acutely and months after TBI, yet little is known about DNAm patterns in pediatric TBI. Given its responsiveness to environmental and biological cues, we hypothesize that DNAm changes occur in children with TBI, even when measured peripherally, reflecting early biological responses to brain injury. Such injury-related epigenetic differences may have relevance for long-term recovery trajectories.

To perform an initial investigation of injury-related DNAm changes, we conducted the Epigenetic Effects on TBI Recovery (EETR) study—a prospective, longitudinal cohort study of children with complicated mild to severe TBI and a comparison group of children with orthopedic injury (OI) and no evidence of brain injury.^25^ As one of the first DNAm studies in complicated mild to severe pediatric TBI, we focused on the brain-derived neurotrophic factor (*BDNF*) gene, a neurotrophin well recognized to be involved in neuronal survival, differentiation, and synaptic strengthening.^26,27^ BDNF levels are upregulated after TBI in preclinical^28–30^ and clinical^31–34^ studies and are associated with mortality and functional outcomes in children^34,35^ and adults^31,36,37^, making it an ideal first target.

This initial report from the EETR study aims to characterize differences in peripheral blood *BDNF* DNAm between children with TBI and OI acutely and at 6- and 12-months post-injury, providing a foundation for future investigation of DNAm patterns in relation to recovery from pediatric TBI.

## 2.0 METHODS

### 2.1 Study overview, design, setting, and participants

The EETR study is an ongoing prospective, longitudinal cohort study of children with complicated mild to severe non-penetrating TBI and a comparison group of children with OI and no evidence of brain injury, conducted at the UPMC Children’s Hospital of Pittsburgh since 2017.^25^ Eligible children are aged 3-18 years and hospitalized at least overnight for their injuries. Complicated mild TBI is defined as a lowest post-resuscitation Glasgow Coma Scale (GCS) score^38^ of 13-15 with clinical neuroimaging indicating intracranial injury; moderate TBI is defined as GCS score of 9-12; severe TBI is defined as GCS score of 3-8. The OI group includes children with bone fractures, excluding skull or facial fractures, and no signs of head trauma or brain injury (nausea/vomiting, headache, loss of consciousness, GCS score <15 at any point) or any suspicion of TBI in the electronic medical record.

Exclusion criteria for both groups includes: (a) non-English-speaking child or parent/guardian (henceforth referred to as parent); (b) documented or parent-reported history of previous TBI or concussion requiring overnight hospitalization; (c) pre-injury neurological disorder or intellectual disability; (d) pre-injury psychiatric disorder requiring hospitalization; (e) spinal cord injury; (f) sensory or motor impairment precluding study measure completion; and (g) pregnancy at time of enrollment.

Additionally, from the beginning of the study until 2/28/2023, children could provide either a blood or saliva biospecimen within 7 days post-injury for study eligibility. Since that date, provision of a blood specimen within 7 days post-injury is required for study eligibility.

Trained research staff perform assessments at three time points, acutely (prior to hospital discharge) and at 6- and 12-months post-injury. Phenotypic data (i.e., clinical, demographic, social factors, etc.) related to the injury, recovery from injury, and/or DNAm are collected. Peripheral blood samples are collected and centrifuged, and the cellular pellet and supernatant are stored at -80°. For families open to donating a blood sample but not to fully participating in the research study, we permit “sample-only” participation. As a result, ∼6% of participants contribute biospecimens without completing questionnaires, which limits the availability of phenotypic data that cannot be extracted from the medical record.

Informed written consent is obtained from parents of all participants, and written assent from children over 8 years of age capable of understanding study procedures. The study is approved by the Institutional Review Board of the University of Pittsburgh (STUDY19040402). A detailed description of the EETR protocol is published.^25^

### 2.2 Covariates

We assessed several *a priori* selected factors previously associated with TBI and/or *BDNF* DNAm (Methods S1). Primary covariates included age, sex, and race (White vs. All other racial groups, combined due to small cell sizes), with race parent-reported and rarely (<2%) supplemented from the medical record. Secondary covariates included age-adjusted body mass index (BMI) z-score (calculated using World Health Organization growth standards^39^); non-head Injury Severity Score (ISS, calculated using a combination of Abbreviated Injury Scale (AIS) and excluding AIS codes related to head injuries^40^); pubertal status (measured using the Pubertal Development Scale^41^); socioeconomic status (SES, composite z-score based on maternal education and census tract income^42^); and psychosocial adversity assessed using the Psychosocial Assessment Tool (PAT)^43^.

### 2.3 *BDNF* DNA methylation data

DNA was extracted from stored peripheral blood cellular pellets using the QIAmp Midi kit (Catalog #51185) from Qiagen Corp (Qiagen, Valencia, CA, USA) and sent to the Center for Inherited Disease Research (CIDR) at Johns Hopkins University (Genetic Resources Core Facility, RRID:SCR_018669) for DNAm profiling using pyrosequencing. Using 500 ng of DNA, concentration and quality checks were completed and bisulfite conversion was performed using standard protocols (Methods S2).

DNAm profiling focused on two *BDNF* CpG sites (i.e., DNAm sites) (Chromosome 11, UCSC Genome Browser hg38 positions 27722033 and 27701612) selected based on a systematic review of epigenetic modifications associated with brain-related phenotypes in humans.^13^ Probe sequences were designed to capture the two target sites and surrounding regions (Table S1). For position 27722033, five sites were captured, and for position 27701612, two sites were captured, resulting in DNAm data for seven DNAm sites in total (Table S2).

### 2.4 Statistical analyses

Analyses were conducted using R version 4.2.1.^44^ DNAm data were analyzed as M values, the log2-transformed ratio of methylated to unmethylated probe intensities.^45^ Descriptive statistics and data checks were conducted by measurement level. Data distributions, patterns, and associations were assessed across all variables. Outliers were identified using the interquartile range (IQR) method, winsorized (extreme ≥3×IQR), and assessed in sensitivity analyses.^46^ Differences in participant characteristics based on DNAm availability were examined to evaluate potential attrition bias. DNAm data were plotted over time. Expanded statistical details are included in the in the Supplementary Information (Methods S3).

Associations between site-specific *BDNF* DNAm and injury type (TBI vs. OI) and TBI severity (severe vs. complicated mild/moderate, combined due to their clinical similarity and small sample size of the moderate group) were assessed using multiple linear regression. While linear mixed-effects models were considered to account for the repeated-measures design, the small sample sizes prevented reliable mixed modeling. Instead, cross-sectional analyses at each time point (acute, 6- months, 12-months) were prioritized to preserve robustness while minimizing model complexity. In these models, DNAm served as the outcome variable, with injury type or injury severity as the primary predictor. Primary models included covariates of age, sex, and race. In sensitivity models, secondary covariates BMI-z, non-head ISS, puberty, SES, and PAT were first added individually to the primary model to assess their influence, and then included together in a final fully adjusted model. For each model, we reported the estimated regression coefficients 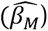, representing the difference in DNAm between groups on the M-value scale, the corresponding 95% confidence intervals (CIs), and adjusted R^2^ values. We also reported the estimated change in DNAm on the Beta-value scale 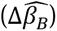 (which ranges from 0 to 100%), calculated by transforming 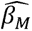 back to the biologically interpretable Beta-value scale and multiplying by 100.^47^

Associations with p-values <0.05 were considered nominally significant. We corrected for multiple testing based on the correlation structure among DNAm sites (Methods S3), resulting in a statistical significance threshold of p<0.0125.

## 3.0 RESULTS

### 3.1 DNAm data cleaning and quality control

Following quality control (Table S2, Figure S1), five of the seven *BDNF* DNAm sites were retained for analyses. One unreliable beta value of zero was set to missing, and thirteen extreme outliers were winsorized to the nearest valid value (Table S3). DNAm data were available at *any* time point from 294 unique participants (189 TBI; 105 OI). Among these, 271 participants (178 TBI; 93 OI) had DNAm data for the acute period; 121 participants (85 TBI; 36 OI) had DNAm data at 6 months; and 103 participants (73 TBI; 30 OI) had DNAm data at 12 months (though it should be noted that site-specific counts varied). Sixty-five participants (51 TBI; 14 OI) had DNAm data available at all three time points for at least one DNAm site. Additional sample size details are presented in the Supplementary Information (Table S4).

### 3.1 Participant characteristics and descriptive overview

Baseline characteristics (collected at the acute time point) are presented in Table 1 for participants with DNAm data available at *any* of the three time points. Participants were 66.3% male, 83.2% White, and had a mean age of 10.6 (±4.4) years at the time of enrollment. Among the TBI group, 58.7% of injuries were classified as complicated mild and 30.2% as severe. No significant differences were observed between injury groups in age, sex, race, BMI-z, SES, PAT, or puberty scores. Non-head ISS was higher in the OI vs. TBI group, consistent with orthopedic injuries affecting body regions outside the head.

**Table 1.** Baseline characteristics of EETR participants with DNA methylation data at any timepoint by injury type.

|  | **TBI**<br>**(n=189)** | **OI**<br>**(n=105)** | **p** |
| --- | --- | --- | --- |
| ***Primary covariates*** |  |  |  |
| Age (years), mean (SD) | 11.77 (4.16) | 11.46 (4.18) | 0.629<sup>a</sup> |
| Sex, n (%) |  |  |  |
| *Male* | 121 (64.0) | 74 (70.5) | 0.321<sup>b</sup> |
| *Female* | 68 (36.0) | 31 (29.5) |  |
| Race (%) |  |  |  |
| *American Indian or Alaskan Native* | 2 (1.1) | 0 | 0.633<sup>c</sup> |
| *Asian* | 2 (1.1) | 2 (1.9) |  |
| *Black or African American* | 21 (11.1) | 11 (10.5) |  |
| *Native Hawaiian or Pacific Islander* | 1 (0.5) | 0 |  |
| *White* | 156 (82.5) | 84 (80.0) |  |
| *Other* | 6 (3.2) | 8 (7.6) |  |
| *Missing, n (%)* | 1 (0.5) | 0 |  |
| ***TBI severity and secondary covariates*** |  |  |  |
| TBI severity, n (%) |  |  |  |
| *Complicated mild* | 111 (58.7) |  |  |
| *Moderate* | 21 (11.1) | NA | NA |
| *Severe* | 57 (30.2) |  |  |
| BMI-z, mean (SD) | 0.56 (1.63) | 0.73 (1.63) | 0.396<sup>a</sup> |
| *Missing* | 1 (0.5) | 6 (5.7) |  |
| Non-head ISS, median [IQR] | 1.0 [1.0, 5.0] | 4.0 [4.0, 9.0] | **0.0001<sup>d</sup>** |
| SES, mean (SD) | -0.03 (0.79) | 0.00 (0.75) | 0.775<sup>a</sup> |
| *Missing, n (%)* | 23 (12.1) | 3 (2.9) |  |
| PAT, median [IQR] | 0.64 [0.30, 1.12] | 0.60 [0.27, 1.17] | 0.832<sup>d</sup> |
| *Missing, n (%)* | 80 (42.1) | 50 (47.6) |  |
| Puberty, mean (SD) | 1.73 (0.95) | 1.76 (1.02) | 0.777<sup>a</sup> |
| *Missing, n (%)* | 76 (40.0) | 41 (39.0) |  |

This table summarizes the characteristics of participants with DNA methylation data available at any of the measured time points (Acute (at injury), 6-months post-injury, or 12-months post-injury).

Expanded participant characteristics are provided in the Supplementary Information (Table S5, Table S6, and Figure S2). Baseline characteristics were similar across groups regardless of DNAm data availability, and across time points within the DNAm subsample (Table S5). Participants with and without DNAm data did not differ on any measures, except for lower SES in the OI group with DNAm data compared to those without DNAm data (p=0.046; Table S6). Within the subsample of participants with DNAm, participants with TBI were more likely to complete the 12 month follow up than those with OI (p=0.0002; Figure S2). There were no statistically significant differences in time post-injury between injury groups as shown in Table 2.

**Table 2.** Time-specific sample sizes and time post-injury of blood samples used for DNA methylation data collection.

| Visit | All |  | TBI |  | OI |  | p<sup>a</sup> |
| --- | --- | --- | --- | --- | --- | --- | --- |
|  | n | Time since injury, mean (SD) | n | Time since injury, mean (SD) | n | Time since injury, mean (SD) |  |
| A | 271 | 31.6 (23.1) hours | 178 | 32.5 (24.9) hours | 93 | 29.8 (19.2) hours | 0.559 |
| 6M | 121 | 216.9 (41.8) days | 85 | 218.0 (38.9) days | 36 | 214.3 (48.6) days | 0.658 |
| 12M | 103 | 405.9 (39.9) days | 73 | 410.4 (39.7) days | 30 | 394.9 (38.6) days | 0.073 |

TBI=traumatic brain injury; OI=orthopedic injury; A=acute (at injury); 6M=6-months post-injury; 12M=12-months post-injury; SD=standard deviation; <sup>a</sup>t-test comparing TBI vs. OI groups.

As part of data cleaning, we assessed covariate missingness (Figure S3) which was most common for PAT, follow-up BMI, and acute puberty assessments, primarily due to mid-study protocol changes and optional measures. As missingness was logistical rather than systematic, complete-case analysis was used for primary models.

In exploring the data, the strongest age-related trends were with puberty and sex (Figure S4). Unadjusted exploratory plots (Figures 1 and S5-S7) showed DNAm differences by injury type (Figure 1) as well as some sex-related variation (Figure S6), with no clear patterns by severity (Figure S5) or race (Figure S7). A heatmap of DNAm-covariate correlations (Figure S8) showed within-DNAm site correlations, but limited associations amongst covariates. Expected links among social factors were observed (Figure S9). A spaghetti plot grouped by injury type illustrates individual variability and DNAm distribution over time (Figure S10).

**Figure 1.**
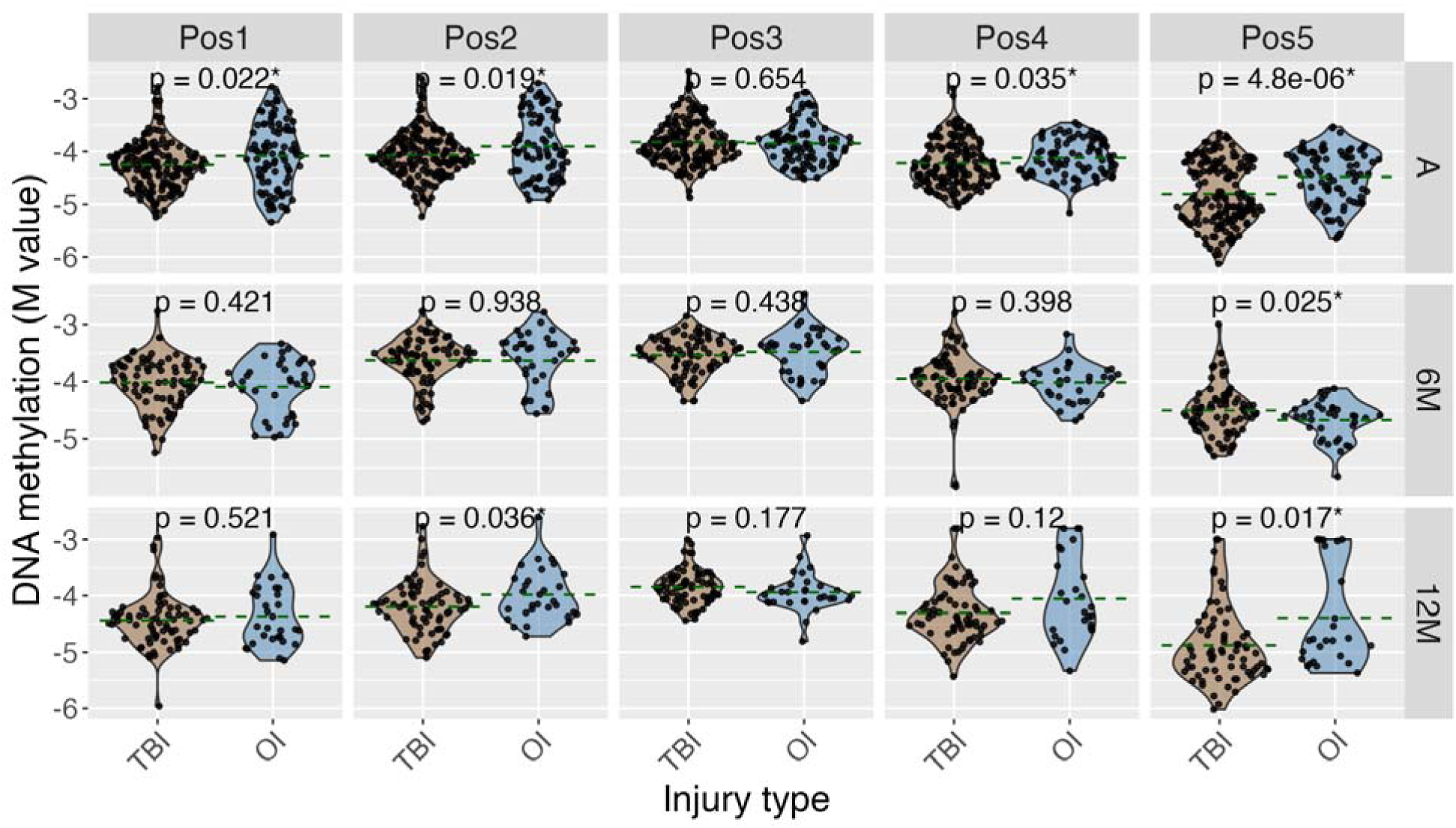
Unadjusted *BDNF* DNA methylation by injury type across time. TBI=traumatic brain injury; OI=orthopedic injury; A=acute (at injury); 6M=6-months post-injury; 12M=12-months post-injury; Pos1-5 indicate site-specific DNAm where Pos1=Position 1 (hg38 chr11:27722033); Pos2=Position 2 (chr11:27722036); Pos3=Position 3 (chr11:27722047); Pos4=Position 4 (chr11:27701612); Pos5=Position 5 (chr11:27701614). Green, horizontal, dashed line indicates group-specific mean values. P-values calculated using t- test (no adjustment for covariates); *p<0.05; refer to Table 3 and Table S7 for association results between *BDNF* DNA methylation and injury type while controlling for covariates.

**Table 3.** Results of linear regression examining associations between injury type (primary predictor, TBI vs. OI as the reference group) and *BDNF* DNAm (outcome) while controlling for age, sex, and race.

| Time | Site | Position (hg38) | n | $\widehat{\beta}_M$ | 95% CI | | *p* | $\Delta\widehat{\beta}_B$ | $R^2_{adj}$ |
| --- | --- | --- | --- | --- | --- | --- | --- | --- | --- |
|  |  |  |  |  | Low | High |  |  |  |
| A | Pos1 | 27722033 | 270 | -0.184 | -0.313 | -0.054 | **0.0060** | -3.18 | 2.98 |
|  | Pos2 | 27722036 | 270 | -0.183 | -0.308 | -0.058 | **0.0044** | -3.17 | 3.57 |
|  | Pos3 | 27722047 | 268 | 0.016 | -0.084 | 0.117 | 0.7510 | 0.28 | -0.27 |
|  | Pos4 | 27701612 | 263 | -0.108 | -0.210 | -0.006 | <u>0.0383</u> | -1.87 | 0.49 |
|  | Pos5 | 27701614 | 265 | -0.338 | -0.482 | -0.194 | **6.48E-06** | -5.83 | 6.56 |
| 6M | Pos1 | 27722033 | 121 | 0.082 | -0.098 | 0.261 | 0.3748 | 1.41 | -1.27 |
|  | Pos2 | 27722036 | 121 | -0.004 | -0.174 | 0.166 | 0.9666 | -0.06 | -2.23 |
|  | Pos3 | 27722047 | 121 | -0.038 | -0.173 | 0.098 | 0.5871 | -0.65 | -0.03 |
|  | Pos4 | 27701612 | 116 | 0.054 | -0.124 | 0.232 | 0.5517 | 0.94 | 0.13 |
|  | Pos5 | 27701614 | 120 | 0.170 | 0.004 | 0.336 | <u>0.0476</u> | 2.94 | 2.20 |
| 12M | Pos1 | 27722033 | 102 | -0.054 | -0.259 | 0.152 | 0.6100 | -0.93 | 1.91 |
|  | Pos2 | 27722036 | 102 | -0.202 | -0.397 | -0.007 | <u>0.0448</u> | -3.50 | 3.21 |
|  | Pos3 | 27722047 | 102 | 0.096 | -0.037 | 0.230 | 0.1601 | 1.67 | -1.11 |
|  | Pos4 | 27701612 | 95 | -0.255 | -0.520 | 0.009 | 0.0620 | -4.42 | 0.14 |
|  | Pos5 | 27701614 | 97 | -0.499 | -0.835 | -0.163 | **0.0045** | -8.56 | 5.21 |

### 3.2 Regression results

Results of linear regression examining associations between injury type (TBI vs. OI; primary predictor) and *BDNF* DNAm values (outcome) while adjusting for primary covariates of age, sex, and race are presented in Table 3. In the acute period, injury type was significantly associated with DNAm at three of the five DNAm sites (positions 1, 2, and 5), with effect sizes 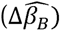 ranging from -3.2 to -5.8, indicating lower *BDNF* DNAm in the TBI group compared to the OI group (p=0.006 to p=6.48E-06). At 12 months post-injury, injury type was significantly associated with DNAm at a single site (position 5, p=0.0045) with the direction of effect consistent with the acute period. No statistically significant associations were observed at the 6-month follow-up.

Results of sensitivity analyses examining associations between injury type and *BDNF* DNAm values while adjusting for primary covariates (age, sex, race) as well as secondary covariates (BMI z-score, ISS, puberty, SES, and/or PAT) resulted in largely consistent results, with injury type significantly associated with lower DNAm at several sites during the acute period, particularly positions 1, 2, 4, and 5 (Table S7). At 6- and 12-months post-injury, few associations reached nominal significance; after correction for multiple testing, none remained significant at 6 months, and only position 5 remained significant at 12 months in select models (Table S7).

Finally, Table 4 shows the results of regression analyses within the TBI subgroup assessing associations between DNAm and injury severity (severe vs. complicated mild/moderate TBI), adjusted for age, sex, and race. No statistically significant differences were observed.

**Table 4.** Results of linear regression examining associations between TBI injury severity (primary predictor, severe vs. complicated mild/moderate (combined) as the reference group) and *BDNF* DNAm (outcome) while controlling for age, sex, and race.

| Time | Position | Position | | $\widehat{\beta}_M$ (*M value*) | 95% CI | | $\Delta\widehat{\beta}_B$ (*Beta, %*) | p |
| --- | --- | --- | --- | --- | --- | --- | --- | --- |
|  |  | (hg38) | n |  | Low | High |  |  |
| A | Pos1 | 27722033 | 177 | 0.080 | -0.066 | 0.227 | 1.39 | 0.2752 |
|  | Pos2 | 27722036 | 177 | 0.041 | -0.099 | 0.182 | 0.72 | 0.5651 |
|  | Pos3 | 27722047 | 175 | 0.036 | -0.092 | 0.163 | 0.62 | 0.5849 |
|  | Pos4 | 27701612 | 173 | 0.017 | -0.121 | 0.155 | 0.29 | 0.8098 |
|  | Pos5 | 27701614 | 175 | 0.034 | -0.159 | 0.226 | 0.59 | 0.7306 |
| 6M | Pos1 | 27722033 | 85 | 0.008 | -0.195 | 0.212 | 0.14 | 0.9360 |
|  | Pos2 | 27722036 | 85 | 0.020 | -0.170 | 0.209 | 0.35 | 0.8373 |
|  | Pos3 | 27722047 | 85 | 0.080 | -0.072 | 0.231 | 1.38 | 0.3063 |
|  | Pos4 | 27701612 | 83 | 0.010 | -0.216 | 0.236 | 0.17 | 0.9313 |
|  | Pos5 | 27701614 | 84 | -0.090 | -0.307 | 0.127 | -1.56 | 0.4187 |
| 12M | Pos1 | 27722033 | 72 | 0.059 | -0.174 | 0.291 | 1.01 | 0.6238 |
|  | Pos2 | 27722036 | 72 | 0.004 | -0.227 | 0.235 | 0.07 | 0.9737 |
|  | Pos3 | 27722047 | 72 | -0.057 | -0.209 | 0.095 | -0.99 | 0.4629 |
|  | Pos4 | 27701612 | 69 | -0.007 | -0.270 | 0.256 | -0.11 | 0.9608 |
|  | Pos5 | 27701614 | 71 | -0.079 | -0.420 | 0.263 | -1.36 | 0.6536 |

TBI=Traumatic brain injury; OI=Orthopedic injury; A=acute (at injury); 6M=6 months post-injury; 12M=12 months post-injury; $\widehat{\beta}_M$ (*M value*)=estimated difference in DNA methylation between participants with severe TBI and those with complicated mild or moderate TBI (reference group), derived from linear regression on the M-value (logit-transformed) scale while holding covariates constant; 95% CI Low / High=lower and upper bounds of 95% confidence interval for $\widehat{\beta}_M$ ; $\Delta\widehat{\beta}_B$ (*Beta, %*)=estimated change in DNA methylation expressed as a percent, derived by transforming $\widehat{\beta}_M$ from the M-value scale to the biologically interpretable Beta value scale and multiplying by 100; Race categorized as White vs. Other races.

## 4.0 DISCUSSION

### 4.1 Key findings

We observed significant differences in *BDNF* DNAm between children with TBI and those with OI at several sites during the acute period (mean of 31.6 hours post-injury) and at one site at 12-months (mean of 405.9 days post-injury). Across all significant findings, DNAm levels were lower in the TBI group (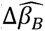 1.87 to 8.56). These differences remained robust across sensitivity models.

### 4.2 Interpretation

These results suggest that *BDNF* DNAm may be responsive to acute injury and may reflect early biological changes that are specific to pediatric TBI, distinct from those observed in children with non-brain injuries. This aligns with early preclinical and adult clinical studies reporting DNAm alterations during the acute and subacute phases of TBI recovery^22,23,48–50^, and supports the broader role of epigenetic regulation in neuroinflammatory processes, stress adaptation, and injury response.^51–54^

While increased DNAm is typically associated with transcriptional repression, the relationship is site- and context-specific.^55^ *BDNF* adds complexity with multiple promoters and non-coding exons that generate transcript isoforms with potentially distinct functions.^56^ In our study, Positions 1-3 lie near Promoter I (exon I), and Positions 4-5 near Promoter IV (exon IV)^13^, suggesting potential differential regulatory relevance in regions linked to brain function and stress response.^57^ Lower *BDNF* DNAm levels observed shortly after TBI may reflect an adaptive response to upregulate expression, consistent with evidence that BDNF protein levels increase following injury.^28–34^ Future work should assess whether DNAm influences isoform-specific expression or protein levels over time, and whether such changes mediate effects of therapeutic or psychosocial interventions.

Although acute group differences were strong and robust across models with different covariates, no significant differences were observed at 6-months, and only one site showed a significant difference at 12-months. The direction of effects remained consistent across all associations, with lower DNAm in the TBI group, but the strength and stability of these associations diminished over time post-injury. This pattern may reflect a transient DNAm response to injury that resolves as recovery progresses.

Alternatively, greater heterogeneity in later recovery stages or reduced sample size may have obscured subtle or sustained effects. Given the complexity of DNAm dynamics and the multifactorial nature of recovery, larger longitudinal studies with additional time points are needed to clarify the significance of these epigenetic changes.

We did not observe associations between *BDNF* DNAm and TBI severity. This may reflect the limited sensitivity of the GCS, especially in pediatric populations^58,59^. Emerging classification systems may provide more biologically meaningful measures of TBI severity.^60^ The observed lack of association could also suggest that factors beyond injury severity (such as psychosocial stressors) play a larger role in DNAm levels post-injury. In fact, adjusted R² values increased when psychosocial adversity (PAT) was added to acute models, despite PAT not showing a significant independent association. This suggests PAT may contribute modestly or interactively to DNAm variability. This merits future work focused on the biological impact of psychosocial adversity following TBI.

Finally, one site (Position 5) exhibited a bimodal distribution in the acute period (Figure 1) that was not explained by covariates, which may reflect unmeasured genetic variation. For example, the *BDNF* Val66Met variant (rs6265)^61,62^ is a well-characterized methylation quantitative trait locus (meQTL)^17,63^ not captured by our study. Genotyping in future studies may help clarify genetic-epigenetic interactions shaping individual response to injury.

### 4.3 Limitations and strengths

The most significant limitation of this study is the reduced sample size at the 6- and 12-month follow-up periods, which limited our ability to include multiple covariates in primary models and precluded the use of more complex longitudinal analyses. Follow-up rates were affected by the COVID- 19 pandemic; the broad geographic catchment area of UPMC Children’s Hospital of Pittsburgh, which made return visits challenging for some families; and participant hesitancy around follow-up blood draws. Importantly though, participants with and without follow-up DNAm data (Figure S2) displayed no significant differences in baseline characteristics, supporting the generalizability of findings. Additional limitations include the single-center nature of the study and the lack of an independent validation cohort, which limit the external validity of our findings. Future multi-site studies with larger, more diverse samples and replication datasets will be important to validate and extend these results.

Despite these limitations, the study has several key strengths. It represents one of the largest pediatric TBI cohorts to date and is strengthened by its prospective design, inclusion of a well-characterized OI comparison group, longitudinal biospecimen collection, and rich phenotypic and epigenetic data. Further, while DNAm varies across tissues and our focus was on brain injury, we intentionally measured DNAm in peripheral blood to identify biomarkers that may reflect systemic responses to injury from an *accessible* and therefore *clinically relevant* tissue—especially important in pediatric populations. Finally, rigorous data cleaning and quality control procedures were implemented, and sensitivity analyses confirmed the robustness of results.

## 5.0 CONCLUSION

This study provides initial evidence of *BDNF* DNAm differences between children with TBI and those with OI, particularly in the acute period, suggesting a role in the brain’s early injury response.

Further research is needed to clarify functional relevance, link DNAm to recovery, explore genome-wide DNAm patterns, and support precision rehabilitation strategies to improve outcomes.

## Data Availability

De-identified DNA methylation and phenotype data will be deposited in dbGaP (accession number phs004096.v1) for participants who provide consent for broad data sharing as part of an NIH R01-funded project currently underway. Until that time, data will be made available upon reasonable request to the Principal Investigator. All requests will be reviewed to ensure consistency with participant consent, institutional policies, and appropriate data use agreements. Please contact Amery Treble-Barna at for more information.

## DECLARATIONS

### Transparency, rigor, and reproducibility summary

The EETR study was pre-registered via publication of a methods paper (https://pubmed.ncbi.nlm.nih.gov/32595586/).^1^ The analysis plan was not formally pre-registered, but the lead analyst (Dr. Lacey Heinsberg) certifies that it was pre-specified.² Sample size was based on availability of participants and biospecimens.³A flow diagram depicting participant screening, eligibility, enrollment, and inclusion in the final analysis is shown in Figure S1. In brief, 954 individuals were screened, 897 were determined to be eligible, 390 were enrolled, and 294 were included in the analyses described here.LJ Biospecimen handling and DNA methylation analyses were performed by laboratory staff blinded to injury group.LJ DNA extracted from peripheral blood was frozen at -80 degrees Celsius between sample processing and data collection. DNA was analyzed in a single batch using research validated pyrosequencing methods and commercially available reagents.LJ All equipment and analytical reagents used to perform measurements on the fluid biomarkers are widely available from commercial sources. Quality control included assessment of replicate agreement, exclusion of unreliable sites, and winsorization of outliers as described.LJ Key inclusion criteria and outcome measures were based on established pediatric TBI diagnostic standards.LJ Statistical assumptions and outliers were analyzed as detailed.^9^ Multiple testing correction was made using a Bonferroni correction based on the correlation structure of the data.¹LJ This is the first report of these findings; replication is planned as part of a larger NIH-funded study (R01NS135492).¹¹ De-identified data will be available through dbGaP (phs004096.v1) for participants who consented to broad sharing. Until then, data may be requested from the PI.¹² Analytic code is available upon request.¹³ All blood samples were investigator- collected, with consent-specific permissions governing any future use.¹LJ The authors agree or have agreed to publish the manuscript using the Mary Ann Liebert Inc. “Open Access” option under appropriate license.¹LJ

### Ethics approval, guidelines, and consent to participate

This study adhered to all ethical considerations, obtaining informed written consent from parents/guardians and child assent for ages 8 years and above. It was approved by the Institutional Review Board of the University of Pittsburgh (STUDY19040402).

### Consent for publication

Not applicable.

### Authors contributions

**Heinsberg:** Conceptualization, Data curation, Formal analysis, Investigation, Methodology, Validation, Visualization, Writing – original draft, Writing – review & editing; **Kesbhat**: Data curation, Project administration, Writing – review & editing; **Petersen**: Data curation, Writing – review & editing; **Kaseman**: Data curation, Project administration, Writing – review & editing; **Stec**: Data curation, Project administration, Writing – review & editing; **Anton**: Data curation, Project administration, Writing – review & editing; **Kochanek**: Writing – review & editing; **Yeates**: Writing – review & editing; **Weeks**: Writing – review & editing; **Conley**: Resources, Writing – review & editing; **Treble-Barna**: Conceptualization, Data curation, Formal analysis, Funding acquisition, Investigation, Methodology, Project administration, Resources, Supervision, Validation, Writing – original draft, Writing – review & editing. All authors contributed to the interpretation of data, critically reviewed the manuscript for important intellectual content, and approved the final version. All authors agree to be accountable for the work.

## Acknowledgements

We thank the participants of this study and their families for generously contributing their time to advance knowledge on pediatric traumatic brain injury recovery.

## Declaration of generative AI and AI-assisted technologies

ChatGPT 4o was used during the development of this manuscript to (1) refine R code for improved data visualization and (2) enhance the clarity, readability, and flow of the written content. All AI-generated output was reviewed, edited, and approved by the authors, who take full responsibility for the final content of the publication.

