## Supplementary Information for "Differential DNA Methylation of the Brain-Derived Neurotrophic Factor Gene is Observed after Pediatric Traumatic Brain Injury Compared to Orthopedic Injury"

**Authors:** Lacey W. Heinsberg*, Aboli Kesbhat; Bailey Petersen,

Lauren Kaseman; Zachary Stec, Nivi Anton, Patrick M. Kochanek,

Keith O. Yeates, Daniel E. Weeks, Yvette P. Conley; Amery Treble-Barna

*Corresponding,

**TABLE OF CONTENTS**

| **Section/Item** | **Description** | **Page** |
| --- | --- | --- |
| **Methods S1** | Expanded covariates. | 2 |
| **Methods S2** | Expanded *BDNF* DNA methylation data collection. | 3 |
| **Table S1** | *BDNF* DNA methylation assay information | 3 |
| **Table S2** | Overview of captured *BDNF* DNA methylation sites and preliminary quality control for longitudinal participant samples. | 3 |
| **Methods S3** | Expanded statistical analyses. | 4 |
| **Figure S1** | Flow diagram of participant screening, eligibility, enrollment, and inclusion in final analysis. | 5 |
| **Table S3** | Summary of site-specific extreme DNA methylation value adjustment. | 6 |
| **Table S4** | Availability of *BDNF* DNA methylation data across sites, time points and time point combinations, and by injury type. | 7 |
| **Table S5** | Expanded baseline characteristics of all EETR participants and the subset with DNA methylation data at Acute (A), 6-month (6M), and 12-month (12M) time points. | 8 |
| **Table S6** | Comparison of baseline characteristics for participants with and without DNA methylation data *at the acute timepoint*, stratified by injury type. | 9 |
| **Figure S2** | Comparison of baseline characteristics by 12-month follow-up status among participants with DNA methylation data at the acute timepoint (A)*.* | 10 |
| **Figure S3** | Missing data patterns across time points for EETR participants with DNA methylation data at any timepoint. | 11 |
| **Figure S4** | Visualization of age distribution and relationships with primary and secondary covariates at baseline in participants with DNA methylation data at the acute timepoint. | 12 |
| **Figure S5** | *BDNF* DNA methylation by TBI injury severity across time. | 13 |
| **Figure S6** | *BDNF* DNA methylation by sex across time. | 14 |
| **Figure S7** | *BDNF* DNA methylation levels by race across time. | 15 |
| **Figure S8** | Heatmap of correlations between *BDNF* DNA methylation and age, age-adjusted BMI z-score, bodily injury severity score, puberty score, socioeconomic proxy measure, and psychosocial adversity. | 16 |
| **Figure S9** | Associations among social drivers of health: socioeconomic proxy measure, psychosocial adversity, and race. | 17 |
| **Figure S10** | Spaghetti plots of *BDNF* DNA methylation over time by injury group. | 18 |
| **Table S7** | Post hoc results of linear regression examining associations between injury type (TBI vs. OI, primary predictor) and *BDNF* DNAm (M values, outcome) while controlling for covariates. | 19 |

**Methods S1.** Expanded covariates.

Age at time of injury and follow up were calculated based on the participant’s date of birth, which was extracted from the medical record along with participant sex. Race was parent-reported and categorized as *White* versus *Other race* due to limited representation of non-White groups, reflecting the demographic composition of Pittsburgh, PA, and to ensure sufficient analytical power.

Height and weight data were extracted from medical record at the time of injury and directly measured by study staff during in-person follow up visits. Age-adjusted BMI z-scores were computed using World Health Organization (WHO) growth standards.^1^

To examine the role of non-head bodily injury severity, we calculated the Injury Severity Score (ISS)^2^ using data from the local trauma registry. The ISS is a standardized measure that reflects overall injury severity in individuals with multiple injuries^2^ and is calculated as the sum of the squares of the three highest Abbreviated Injury Scale (AIS) scores across distinct body regions. To isolate non-head injury severity, we excluded AIS codes related to head injuries (i.e., we used the three highest non-head regions). ISS scores range from 1 to 75, with greater scores indicating greater bodily injury severity.

Pubertal status was estimated for children ≥8 years using the self-report Pubertal Development Scale.^3^ The scale provides a puberty score ranging from 1 to 4, with 1 indicating no pubertal development and 4 indicating completed pubertal development, and shows adequate agreement with direct clinical assessment of Tanner staging.^3^ Participants under the age of 8 were assigned a value of 1.

As indicators of social drivers of health, an SES proxy was calculated as a composite z-score, created by standardizing maternal years of education (self-reported by the parent) and the median income for the participant’s census tract (based on their residential address) using the study sample's mean and standard deviation.^4^ Psychosocial adversity was assessed using the Psychosocial Assessment Tool (PAT)^5^, a caregiver-report screening instrument designed to assess psychosocial risk in families of children with medical conditions. The PAT evaluates a variety of domains: family structure and resources, social support, patient, sibling, and caregiver problems, family beliefs, and caregiver stress reactions. We examined the PAT total score, which ranges between 0.00 and 7.00, with higher scores indicating greater psychosocial burden.

**Methods S2.** Expanded *BDNF* DNA methylation data collection.

*BDNF* DNA methylation (DNAm) data were generated at the Center for Inherited Disease Research (CIDR) at Johns Hopkins University (Genetic Resources Core Facility, RRID:SCR_018669). Using 1 μg of DNA, bisulfite conversion was performed with the Qiagen EpiTect Bisulfite kit (Catalog 59104 [48 reactions] or 59110 [96 reactions]). PCR reactions utilized the PyroMark PCR Kit (Catalog 978703 [200 reactions] or 978705 [800 reactions]). When possible, a 1:10 dilution of the bisulfite-converted DNA was prepared, and 6 μL of the dilution was added to the PCR reaction. Standard PCR conditions were followed. Reagents used included: (a) PyroMark Q48 Advanced Reagents (Catalog 974002) or PyroMark Q48 Advanced CpG Reagents (Catalog 974022); (b) PyroMark Q48 Magnetic Beads (Catalog 974203); (c) PyroMark Q48 Discs (Catalog 974901); and (d) PyroMark Q48 Absorber Strips (Catalog 974912).

Target position assays (**Table S1**) were designed by CIDR staff using the Qiagen PyroMark Assay Design Software 2.0. DNAm data were collected via bisulfite pyrosequencing using the Qiagen PyroMark Q48 instrument following the standard low-concentration protocol per the PyroMark Q48 Autoprep User Manual, with manual primer addition. Quality control included water controls and methylation standards ranging from 0% (Catalog D5013-1) to 100% (Catalog D5014). The software’s algorithm automatically assigned “Pass”, “Check”, or “Fail” values to each sample and data were manually reviewed by staff using the Qiagen Q48 Software (version 2.4.2).

**Table S1.** *BDNF* DNA methylation assay information.

|  | **chr11:27722033 (hg38)** | **chr11:27701612 (hg38)** |
| --- | --- | --- |
| Sequence before conversion | **CG**CCGGCAGACTACCGCTTTAATAATAATACCAGAAAAGCGCAGCAGGGAGGGGGTGGGGGGCGGCAA | TC**CG**CGGTGAATGGGAAAGTGGGTGGGA |
| Sequence to analyze | YGTYGGTAGATTATYGTTTTAATAATAATATTAGAAAAGYGTAGTAGG GAGGGGGTGGGGGGYGGTAA | TTYGYGGTGAATGGGAAAGTGGGTGGGA |
| Nucleotide dispensation | GTCAGTCTGCTAGATGATCTGTTATATATATAGAATGTCGTAGTAGAG GTGGTCGT | GTCTAGTCGTGATGAG |
| Number of sites captured | 5 | 2 |

Probe sequences were designed to capture the two target sites (hg38 chr11:27722033 and chr11:27701612) and surrounding regions. For position 27722033, five sites were captured, and for position 27701612, two sites were captured, resulting in DNAm data for seven sites in total. The study included 574 samples: 497 participant samples as well as 14 duplicate participant samples (collected at up to three time points: acute, 6 months, and 12 months post-injury), 16 blank water controls, and 48 methylation standards. Quality control steps involved evaluating the performance of blank water controls and methylation standards (0%, 5%, 25%, 50%, 75%, and 100% methylation, with 8 samples for each category). As expected, blank water controls were not analyzable due to the absence of data. Over 95% of methylation standards were labeled as “Passed” by the Qiagen software, except for two dropped sites. Data collection also included 14 technical duplicates, which showed Pearson correlation coefficients ranging from 0.23 to 0.62 across sites. Control and duplicate samples were removed before analysis.

Lab quality control for the participant samples are shown below (**Table S2**). For quality control, CpG sites that “failed” in more than 5% of participant samples were excluded from the analysis. Any individual samples marked as “Fail” were removed before analyses. Software output was manually inspected for “Check” values and, after quality control and adjustment of extreme outliers (see Table S6), we conducted analyses using only “passed” samples and again using both “passed + check” samples. Results were concordant across both sets, so we present findings from the “passed + check” sample to maximize sample size. After quality control, we had DNAm data for 294 participants *from at least one time point*. As detailed above, “Check” values were included in final analyses after exploring their influence in sensitivity analyses.

**Table S2.** Overview of captured *BDNF* DNA methylation sites and preliminary quality control for longitudinal participant samples.

|  |  | | **chr11: 27722033** | | | | | | | |
| --- | --- | --- | --- | --- | --- | --- | --- | --- | --- | --- |
| **Position (hg38)** | | **Position (hg19)** | | **Illumina cg probe** | **Check** | **Failed** | **Not analyzable** | **Passed** | **Status** | **Label Assigned** |
| 27722033 | | 27743580 | | cg16257091 | 1 (0.2) | 0 | 3 (0.6) | 493 (99.2) | Retained | Pos1 |
| 27722036 | | 27743583 | | N/A | 1 (0.2) | 0 | 3 (0.6) | 493 (99.2) | Retained | Pos2 |
| 27722047 | | 27743594 | | N/A | 16 (3.2) | 2 (0.4) | 3 (0.6) | 476 (95.8) | Retained | Pos3 |
| 27722072 | | 27743619 | | cg03167496 | 396 (79.7) | 90 (18.1) | 3 (0.6) | 8 (1.6) | Dropped | N/A |
| 27722095 | | 27743642 | | N/A | 396 (79.7) | 91 (18.3) | 3 (0.6) | 7 (1.4) | Dropped | N/A |
|  |  | | **chr11: 27701612** | | | | | | | |
| **Position (hg38)** | | **Position (hg19)** | | **Illumina cg probe** | **Check** | **Failed** | **Not analyzable** | **Passed** | **Status** | **Label Assigned** |
| 27701612 | | 27723159 | | N/A | 53 (10.7) | 19 (3.8) | 3 (0.6) | 422 (84.9) | Retained | Pos4 |
| 27701614 | | 27723161 | | N/A | 31 (6.2) | 10 (2.0) | 3 (0.6) | 453 (91.2) | Retained | Pos5 |

Notes: Table presents participant samples only (longitudinal) and excludes methylation standards and duplicates.

**Methods S3.** Expanded statistical analyses.

All statistical analyses and graphics were generated using R version 4.2.1.^6^ DNAm data were analyzed as M values, the log2-transformed ratio of methylated to unmethylated probe intensities, which offer improved statistical properties over beta values.

Descriptive statistics were computed for all variables according to measurement level (counts and frequency percentages for categorical variables and means and standard deviations (SD) or medians and interquartile ranges (IQR) for continuous variables). Data distributions, patterns, and associations were assessed across all variables, including across TBI and OI groups, using graphical and statistical methods. These included chi-square tests for categorical variables, t-tests or Wilcoxon rank-sum tests for two-group comparisons, and One-Way Analysis of Variance (ANOVA) or Kruskal-Wallis tests for comparisons across more than two groups, depending on distributional assumptions. Pearson or Spearman correlation coefficients were calculated as appropriate. Data visualizations included sina/violin plots and scatterplots, generated using the *ggplot2* R package.^7^ Outliers were assessed using the interquartile range (IQR) method, with extreme outliers defined as values beyond 3 times the IQR.^8^ Identified outliers were winsorized and reported, and sensitivity analyses were conducted to evaluate their influence on results.

Given the longitudinal nature of the study, DNAm data were plotted over time to explore potential temporal trends between TBI and OI groups. Differences in participant characteristics based on DNAm data availability (both overall and across study time points) was assessed to examine any potential biases due to attrition.

Associations between site-specific *BDNF* DNAm and injury type (TBI vs. OI) and TBI severity (severe vs. complicated mild/moderate) was assessed using multiple linear regression. While linear mixed-effects models were considered to account for the repeated-measures design, the small sample sizes limited the statistical power necessary for reliable mixed modeling. Instead, cross-sectional analyses at each time point (acute, 6 months, 12 months) were prioritized to preserve robustness while minimizing model complexity.

In these models, DNAm served as the outcome variable, with injury type or injury severity as the primary predictor. Primary models included covariates of age, sex, and race. Secondary covariates (BMI-z, non-head ISS, puberty, SES, and PAT) were first added individually to the primary model to assess their influence, and then included together in a final fully adjusted model. For each model, we reported the estimated regression coefficients $(\hat{\beta_{M}}$) representing the difference in DNAm between groups on the M-value scale, the corresponding 95% confidence intervals (CIs), and adjusted R^2^ values. We also reported the estimated change in DNAm on the Beta-value scale ($\Delta\hat{\beta_{B}}$), calculated by transforming $\hat{\beta_{M}}$ to the biologically interpretable Beta-value scale and multiplying by 100. Associations with p-values <0.05 were considered nominally significant. However, we corrected for multiple testing using the *meff()* function from the *poolr* R package.^9^ Specifically, we calculated the number of effective tests (N_eff_) based on the correlation structure among DNAm sites. A Bonferroni correction was then applied (α=0.05/N_eff_), resulting in a statistical significance threshold of p<0.0125.

**Figure S1.** Flow diagram of participant screening, eligibility, enrollment, and inclusion in final analysis.

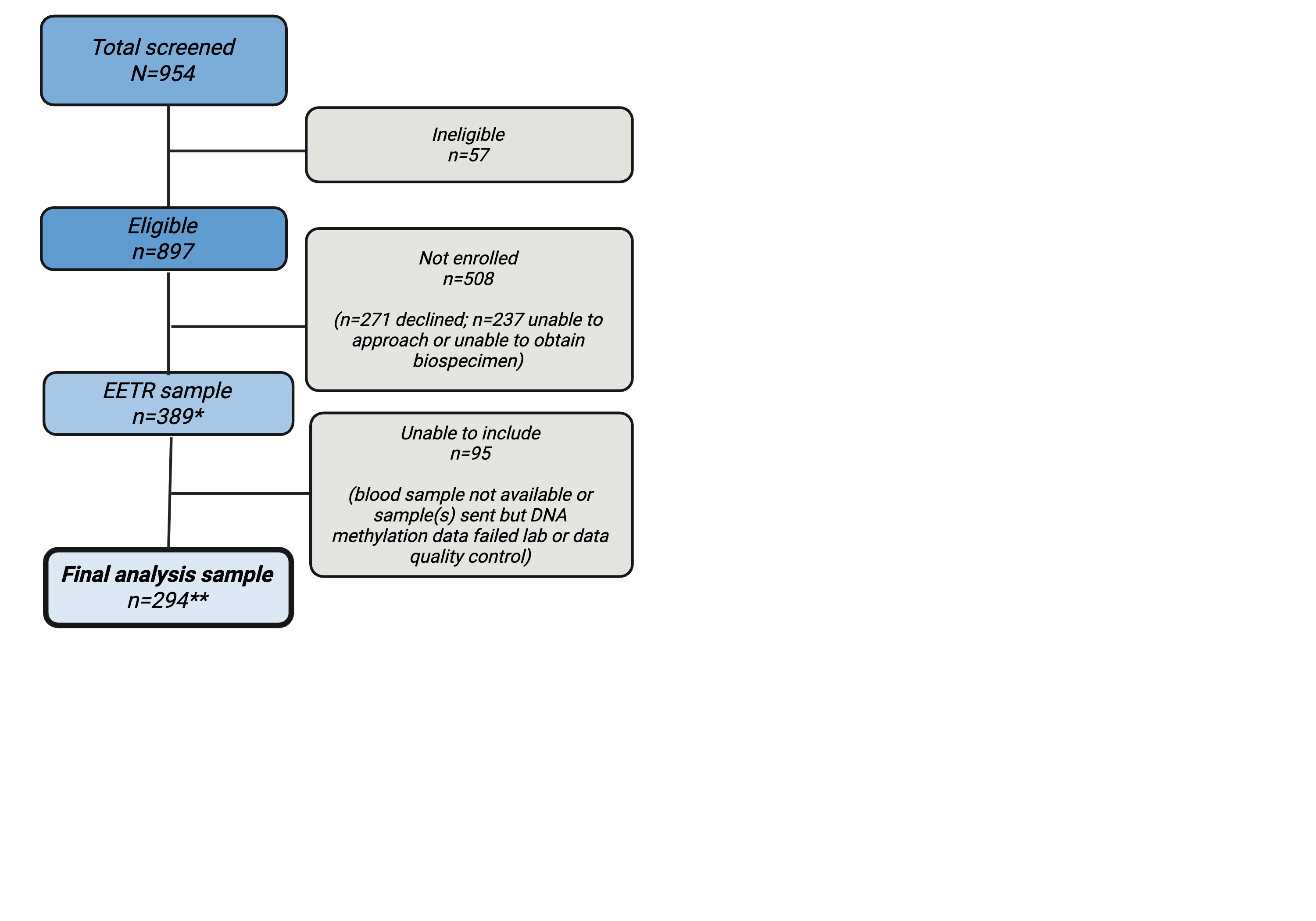

Abbreviations: EETR=Epigenetic Effects on Pediatric Traumatic Brain Injury Recovery. Notes: *EETR recruitment is ongoing (recruitment extended as part of an R01), flow chart represents sample sizes at the time of DNA methylation data collection; n=22 consented for "sample only" (see methods). **Final analysis sample represents the number of participants with DNA methylation data available at *any* time point post-quality control (acute [at injury]; 6-months post-injury; or 12-months post-injury) (see Table S2 for sample-level details).

**Table S3.** Summary of site-specific extreme DNA methylation value adjustment.

| Position | Original | Adjusted |
| --- | --- | --- |
| Pos1 | 19.71 | 12.76 |
| Pos1 | 14.73 | 12.76 |
| Pos1 | 14.73 | 12.76 |
| Pos1 | 14.73 | 12.76 |
| Pos1 | 14.73 | 12.76 |
| Pos3 | 27.41 | 15.25 |
| Pos4 | 14.96 | 12.99 |
| Pos4 | 13.75 | 12.99 |
| Pos4 | 14.15 | 12.99 |
| Pos4 | 13.47 | 12.99 |
| Pos5 | 14.9 | 12.11 |
| Pos5 | 12.46 | 12.11 |
| Pos5 | 12.55 | 12.11 |
| Pos5 | 0* | NA |

Pos1-5 indicate site-specific DNAm where Pos1=Position 1 (hg38 chr11:27722033); Pos2=Position 2 (chr11:27722036); Pos3=Position 3 (chr11:27722047); Pos4=Position 4 (chr11:27701612); Pos5=Position 5 (chr11:27701614). Extreme outliers were adjusted to the next nearest value not considered an outlier based on 3xIQR (interquartile range). *This value was deemed unreliable and removed (set to NA).

**Table S4.** Availability of *BDNF* DNA methylation data across sites, time points and time point combinations, and by injury type.

| **Table S4a1.** Number of participants with DNAm data at each individual time point (site-specific and post-QC counts). | | | | | |
| --- | --- | --- | --- | --- | --- |
| Time | Pos1 | Pos2 | Pos3 | Pos4 | Pos5* |
| A | 271 | 271 | 269 | 264 | 266 |
| 6M | 121 | 121 | 121 | 116 | 120 |
| 12M | 102 | 102 | 102 | 95 | 97 |
| **Table S4a2.** Identical data to S4a1, now stratified by injury type (TBI and OI). | | | | | |
| TBI | | | | | |
| Time | Pos1 | Pos2 | Pos3 | Pos4 | Pos5* |
| A | 178 | 178 | 176 | 174 | 176 |
| 6M | 85 | 85 | 85 | 83 | 84 |
| 12M | 72 | 72 | 72 | 69 | 71 |
| OI | | | | | |
| Time | Pos1 | Pos2 | Pos3 | Pos4 | Pos5* |
| A | 93 | 93 | 93 | 90 | 90 |
| 6M | 36 | 36 | 36 | 33 | 36 |
| 12M | 30 | 30 | 30 | 26 | 26 |
| **Table S4b1.** Number of participants with DNAm data at one or more combinations of time points (e.g., A+6M, A+6M+12M) (site-specific and post-QC counts). | | | | | |
| Time | Pos1 | Pos2 | Pos3 | Pos4 | Pos5* |
| A | 153 | 153 | 153 | 152 | 152 |
| 6M | 4 | 4 | 4 | 10 | 9 |
| 12M | 2 | 2 | 3 | 1 | 1 |
| A + 6M | 35 | 35 | 35 | 31 | 32 |
| A + 12M | 18 | 18 | 17 | 19 | 17 |
| 6M + 12M | 17 | 17 | 18 | 13 | 14 |
| A + 6M + 12M | 65 | 65 | 64 | 62 | 65 |
| **Table S4b2.** Identical data to S4b1, now stratified by injury type (TBI and OI). | | | | | |
| TBI | | | | | |
| Time | Pos1 | Pos2 | Pos3 | Pos4 | Pos5* |
| A | 90 | 90 | 90 | 88 | 90 |
| 6M | 3 | 3 | 3 | 5 | 5 |
| 12M | 1 | 1 | 2 | 1 | 1 |
| A + 6M | 24 | 24 | 24 | 24 | 22 |
| A + 12M | 13 | 13 | 12 | 14 | 13 |
| 6M + 12M | 7 | 7 | 8 | 6 | 6 |
| A + 6M + 12M | 51 | 51 | 50 | 48 | 51 |
| OI | | | | | |
| Time | Pos1 | Pos2 | Pos3 | Pos4 | Pos5* |
| A | 63 | 63 | 63 | 64 | 62 |
| 6M | 1 | 1 | 1 | 5 | 4 |
| 12M | 1 | 1 | 1 | 0 | 0 |
| A + 6M | 11 | 11 | 11 | 7 | 10 |
| A + 12M | 5 | 5 | 5 | 5 | 4 |
| 6M + 12M | 10 | 10 | 10 | 7 | 8 |
| A + 6M + 12M | 14 | 14 | 14 | 14 | 14 |

Abbreviations: TBI=traumatic brain injury; OI=orthopedic injury; A=acute (at injury); 6M=6-months post-injury; 12M=12-months post-injury; Pos1-5 indicate site-specific DNAm where Pos1=Position 1 (hg38 chr11:27722033); Pos2=Position 2 (chr11:27722036); Pos3=Position 3 (chr11:27722047); Pos4=Position 4 (chr11:27701612); Pos5=Position 5 (chr11:27701614). Notes: **Tables S4a1** and **S4a2** show the number of participants with available DNA methylation data at each individual time point (A, 6M, 12M) for the five sites, regardless of data availability at other time points. Participant counts reflect data availability at each site and/or time point and do not necessarily represent the same individuals across sites or time points. As a result, totals in other tables (e.g., Table 2, Table S5) may differ slightly due to the inclusion of non-overlapping samples. **Tables S4b1** and **S4b2** show complete-case counts for time point combinations, meaning participants are only included if they had data at *all* listed time points for a given site (e.g., A+6M+12M requires data at all three time points). *****One zero value at Pos5 was determined to be an artifact and set to missing (NA, Table S3); as a result, counts for Pos5 in this table differ by one from those reported in Table S2.

**Table S5.** Expanded baseline characteristics of all EETR participants and the subset with DNA methylation data at Acute (A), 6 month (6M), and 12 month (12M) time points.

| **Table S5a.** Time-varying variables. | | | | | | | | |
| --- | --- | --- | --- | --- | --- | --- | --- | --- |
|  | **All of EETR** | | **EETR *BDNF* DNA methylation subsample** | | | | | |
|  | **Acute** | | **Acute** | | **6M** | | **12M** | |
|  | **TBI (n=234)** | **OI (n=155)** | **TBI (n=178)** | **OI (n=93)** | **TBI (n=85)** | **OI (n=36)** | **TBI (n=73)** | **OI (n=30)** |
| Age (years), mean (SD) | 10.48 (4.44) | 10.93 (4.29) | 10.68 (4.40) | 10.56 (4.32) | 11.48 (4.35) | 11.67 (4.41) | 11.96 (4.31) | 11.29 (4.29) |
| BMI (kg/m2), mean (SD) | 19.76 (5.47) | 20.28 (5.48) | 19.68 (5.34) | 20.34 (6.15) | 20.06 (4.18) | 21.80 (7.03) | 19.83 (4.54) | 23.86 (9.30) |
| *Missing, n (%)* | 2 (0.85) | 13 (8.4) | 1 (0.6) | 6 (6.5) | 19 (22.4) | 15 (41.7) | 12 (16.4) | 14 (46.7) |
| BMI-z, mean (SD) | 0.56 (1.69) | 0.78 (1.52) | 0.51 (1.63) | 0.79 (1.63) | 0.56 (1.31) | 0.85 (1.59) | 0.43 (1.17) | 1.30 (1.88) |
| *Missing, n (%)* | 2 (0.85) | 13 (8.4) | 1 (0.6) | 6 (6.5) | 19 (22.4) | 15 (41.7) | 12 (16.7) | 14 (46.7) |
| PAT, median (IQR) | 0.62 [0.30, 1.10] | 0.59 [0.26, 1.14] | 0.63 [0.30, 1.12] | 0.59 [0.26, 1.18] | 0.74 [0.34, 1.28] | 0.46 [0.24, 0.84] | 0.74 [0.35, 1.08] | 0.52 [0.17, 0.92] |
| *Missing, n (%)* | 74 (31.6) | 58 (37.4) | 61 (34.3) | 39 (41.9) | 12 (14.1) | 1 (2.8) | 13 (18.1) | 2 (6.7) |
| Time since injury for DNAm measure, mean (SD) | NA | | 32.55 (24.91) | 29.75 (19.22) | 218.05 (38.86) | 214.25 (48.61) | 410.37 (39.78) | 394.90 (38.57) |
| **Table S5b.** Static variables. | | | | | | | | |
|  | **All of EETR** | | **EETR *BDNF* DNA methylation subsample** | | | | | |
|  | **Acute** | | **Acute** | | **6M** | | **12M** | |
|  | **TBI (n=234)** | **OI (n=155)** | **TBI (n=178)** | **OI (n=93)** | **TBI (n=85)** | **OI (n=36)** | **TBI (n=73)** | **OI (n=30)** |
| Sex, n (%) |  |  |  |  |  |  |  |  |
| *Male* | 149 (63.7) | 109 (70.3) | 114 (64.0) | 66 (71.0) | 53 (62.4) | 29 (80.6) | 46 (63.0) | 21 (70.0) |
| *Female* | 85 (36.3) | 46 (29.7) | 64 (36.0) | 27 (29.0) | 32 (37.6) | 7 (19.4) | 27 (37.0) | 9 (30.0) |
| Race (%), n (%) |  |  |  |  |  |  |  |  |
| *White* | 195 (83.3) | 128 (82.5) | 148 (83.1) | 74 (79.6) | 73 (85.9) | 27 (75.0) | 60 (82.2) | 25 (83.3) |
| *Other race* | 38 (16.2) | 27 (17.5) | 29 (16.3) | 19 (20.4) | 12 (14.1) | 9 (25.0) | 13 (17.8) | 5 (16.7) |
| *Missing* | 1 (0.4) | 0 | 1 (0.6) | 0 | 0 | 0 | 0 | 0 |
| TBI severity, n (%) |  |  |  |  |  |  |  |  |
| *Complicated mild* | 136 (58.1) | NA | 105 (59.0) | NA | 50 (58.8) | NA | 40 (54.8) | NA |
| *Moderate* | 26 (11.1) |  | 19 (10.7) |  | 11 (12.9) |  | 10 (13.7) |  |
| *Severe* | 72 (30.8) |  | 54 (30.3) |  | 24 (28.2) |  | 23 (31.5) |  |
| Non-head ISS, median [IQR] | 1.0 [1.0, 5.0] | 4.0 [4.0, 5.5] | 1.0 [1.0, 5,0] | 4.0 [4.0, 9.0] | 1.0 [1.0, 5.0] | 4.0 [4.0, 6.0] | 1.0 [1.0, 5.0] | 4.0 [4.0, 5.0] |
| SES, mean (SD) | -0.03 (0.78) | 0.05 (0.77) | 0.0 (0.77) | -0.05 (0.74) | -0.02 (0.84) | 0.26 (0.77) | -0.02 (0.76) | 0.23 (0.83) |
| *Missing, n (%)* | 28 (11.9) | 5 (3.2) | 23 (12.9) | 3 (3.2) | 1 (1.2) | 0 | 1 (1.4) | 0 |
| Puberty, median [IQR] | 1.67 (0.93) | 1.86 (1.04) | 1.73 (0.95) | 1.78 (1.0) | 2.13 (1.13) | 2.02 (0.99) | 2.28 (1.09) | 2.20 (1.08) |
| *Missing, n (%)* | 92 (39.1) | 62 (40.0) | 73 (41.0) | 34 (36.6) | 10 (11.8) | 1 (2.78) | 5 (6.9) | 1 (3.33) |

Group definitions: **All of EETR**=the full baseline cohort of EETR participants (TBI and OI) with available baseline (acute, Time A) data, regardless of biospecimen availability; **EETR BDNF DNA methylation subsample**=subsets of participants who had DNA methylation data available at the acute (A), 6-month (6M), or 12-month (12M) timepoints. In all columns, the table summarizes baseline (acute timepoint) characteristics of these groups, meaning the 6M and 12M columns reflect participants who contributed methylation data at those future timepoints, but their baseline data is shown here. Therefore, participant counts may not match other tables exactly. Abbreviations: TBI=traumatic brain injury; OI=orthopedic injury; A=acute (at injury); 6M=6-months post-injury; 12M=12-months post-injury; DNAm=DNA methylation; BMI=body mass index (kg/m²); BMI-z=age-adjusted body mass index Z score (World Health Organization growth standards); PAT=psychosocial assessment tool total score; ISS=non-head injury severity score; SES=socioeconomic proxy score; Puberty=puberty score; SD=standard deviation; IQR=interquartile range. Notes: Time since injury for DNAm measure=number of hours (A) or days (6M and 12M) since injury at the time of DNAm assessment.

**Table S6.** Comparison of baseline characteristics for participants with and without DNA methylation data *at the acute timepoint*, stratified by injury type.

|  | **TBI, DNAm (n=178)** | **TBI, no DNAm (n=56)** | ***p*** | **OI, DNAm**  **(n=93)** | **OI, no DNAm**  **(n=62)** | ***p*** |
| --- | --- | --- | --- | --- | --- | --- |
| ***Primary covariates*** | | | | | | |
| Age (years), mean (SD) | 10.68 (4.40) | 9.84 (4.55) | 0.218 | 10.56 (4.32) | 11.49 (4.22) | 0.187 |
| Sex, n (%) |  |  |  |  |  |  |
| *Male* | 114 (64.0) | 35 (62.5) | 0.833 | 66 (71.0) | 43 (69.4) | 0.830 |
| *Female* | 64 (36.0) | 21 (37.5) |  | 27 (29.0) | 19 (30.6) |  |
| Race (%), n (%) |  |  |  |  |  |  |
| *White* | 148 (83.1) | 47 (83.9) | 0.956 | 74 (79.6) | 54 (87.1) | 0.226 |
| *Other race* | 29 (16.3) | 9 (16.1) |  | 19 (20.4) | 8 (12.9) |  |
| *Missing* | 1 (0.6) | 0 |  | 0 | 0 |  |
| ***Secondary covariates and other participant characteristics*** | | | | | | |
| BMI (kg/m^2^), mean (SD) | 19.68 (5.34) | 20.01 (5.94) | 0.695 | 20.34 (6.15) | 20.20 (4.28) | 0.876 |
| *Missing, n (%)* | 1 (0.6) | 1 (1.8) |  | 6 (6.5) | 7 (11.3) |  |
| BMI-z, mean (SD) | 0.51 (1.63) | 0.73 (1.88) | 0.397 | 0.79 (1.63) | 0.76 (1.32) | 0.904 |
| *Missing, n (%)* | 1 (0.6) | 1 (1.8) |  | 6 (6.5) | 7 (11.3) |  |
| TBI severity, n (%) |  |  |  |  |  |  |
| *Complicated mild* | 105 (59.0) | 31 (55.4) | 0.874 | NA | | |
| *Moderate* | 19 (10.7) | 7 (12.5) |  |  |  |  |
| *Severe* | 54 (30.3) | 18 (32.1) |  |  |  |  |
| Non-head ISS, median [IQR] | 1.0 [1.0, 5.0] | 1.0 [1.0, 5.3] | 0.501 | 4.0 [4.0, 9.0] | 4.0 [4.0, 5.0] | 0.373 |
| SES, mean (SD) | 0.0 (0.77) | -0.10 (0.80) | 0.402 | -0.05 (0.74) | 0.20 (0.78) | **0.046** |
| *Missing, n (%)* | 23 (12.9) | 5 (8.9) |  | 3 (3.2) | 2 (3.2) |  |
| PAT, median (IQR) | 0.63 [0.30, 1.12] | 0.61 [0.32, 1.09] | 0.911 | 0.59 [0.26, 1.18] | 0.59 [0.29, 1.06] | 0.962 |
| *Missing, n (%)* | 61 (34.3) | 13 (23.2) |  | 39 (41.9) | 19 (30.6) |  |
| Puberty, median [IQR] | 1.73 (0.95) | 1.51 (0.85) | 0.268 | 1.78 (1.0) | 2.0 (1.12) | 0.419 |
| *Missing, n (%)* | 73 (41.0) | 19 (33.9) |  | 34 (36.6) | 28 (45.2) |  |

Group definitions: **TBI or OI, DNAm**=participants with acute blood samples and DNA methylation (DNAm) data available; **TBI or OI, no DNAm**=participants without acute blood samples and DNAm data. Abbreviations: TBI=traumatic brain injury; OI=orthopedic injury; A=acute (at injury); 6M=6-months post-injury; 12M=12-months post-injury; DNAm=DNA methylation; BMI=body mass index (kg/m²); BMI-z=age-adjusted body mass index Z score (World Health Organization growth standards); PAT=psychosocial assessment tool total score; ISS=non-head injury severity score; SES=socioeconomic proxy score; Puberty=puberty score; SD=standard deviation; IQR=interquartile range. Notes: Comparisons were made within each injury type (TBI or OI) using t-tests or Wilcoxon rank-sum tests for continuous variables and chi-square or Fisher’s exact tests for categorical variables, as appropriate.

**Figure S2.** Comparison of baseline characteristics by 12-month follow-up status among participants with DNA methylation data at the acute timepoint (A)*.*

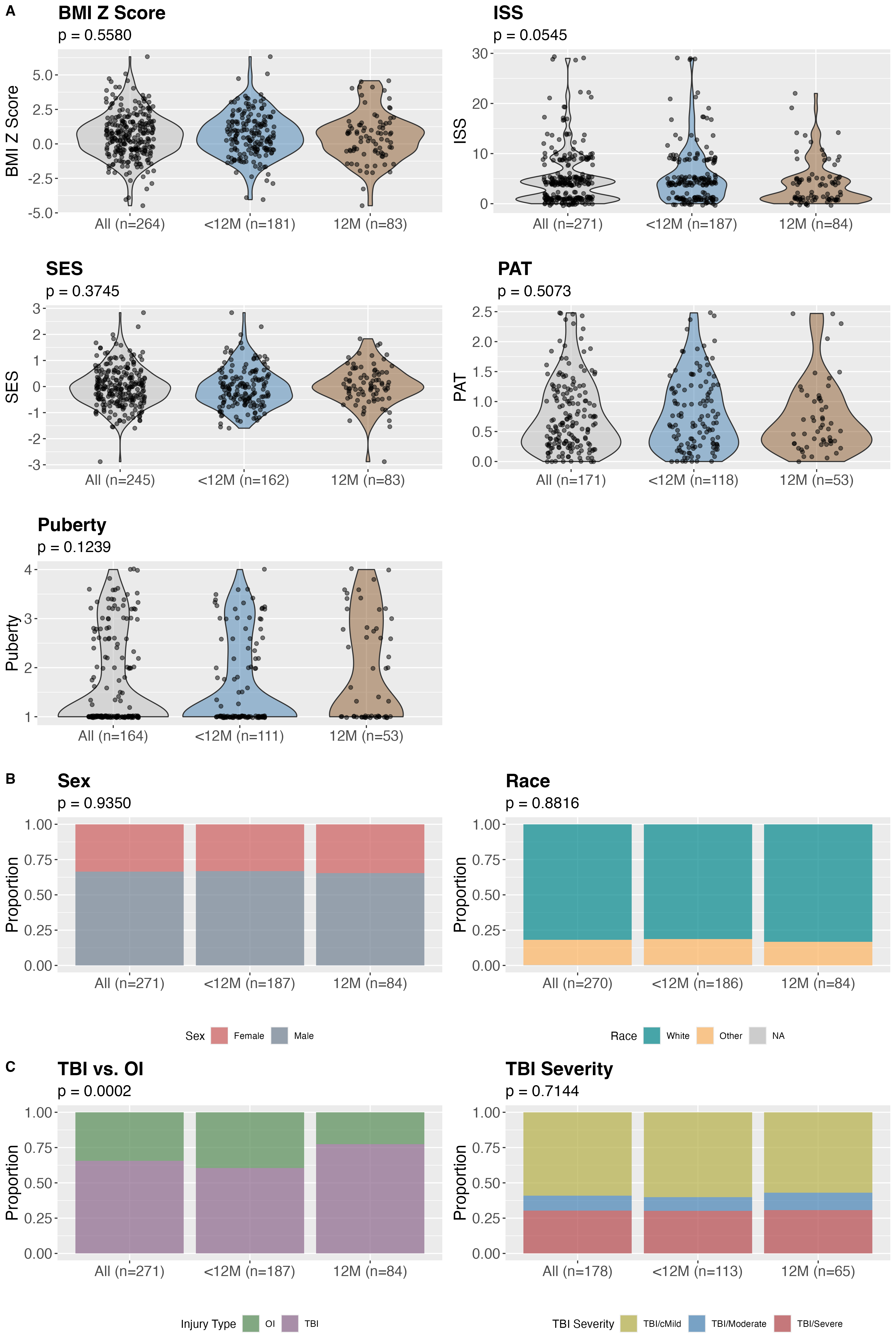

Group definitions: **All**=All participants who had DNAm data available at the acute (A) time point; **<12M**=participants who had DNAm data available at the acute (A) time point but were “lost” to follow-up before 12 months (either due to being lost from the study, not completing an in-person 6M and/or 12M visit, or missing blood collection at those visits); **12M**=participants who had DNAm data available at the acute (A) and 12M time points. Therefore, participant counts may not match other tables exactly. Abbreviations: TBI=traumatic brain injury; OI=orthopedic injury; BMI=body mass index; ISS=non-head injury severity score; SES=socioeconomic proxy score; PAT=psychosocial assessment tool total score; Puberty=puberty score; cMild=complicated mild. Notes: **Figure S1.A.** Raincloud plots of values for continuous variables, including BMI z-score, SES, ISS, PAT, and puberty score. T-tests were used for normally distributed variables (BMI z-score, SES); Wilcoxon rank-sum tests were used for skewed variables (ISS, PAT, and puberty score). **Figure S1.B.** Proportional bar plots for categorical variables (sex, race), with p-values from chi-square tests of independence. **Figure S1.C.** Proportions of injury type (TBI vs. OI) and TBI severity levels (cMild, moderate, severe) within the TBI group by follow-up status, with p-values from chi-square tests. *Statistical comparisons exclude the "All" group; tests compare only "<12M" vs. "12M"; see Table S5 for a comparison of participants with and without DNAm data.*

**Figure S3.** Missing data patterns across time points for EETR participants with DNA methylation data at any timepoint.

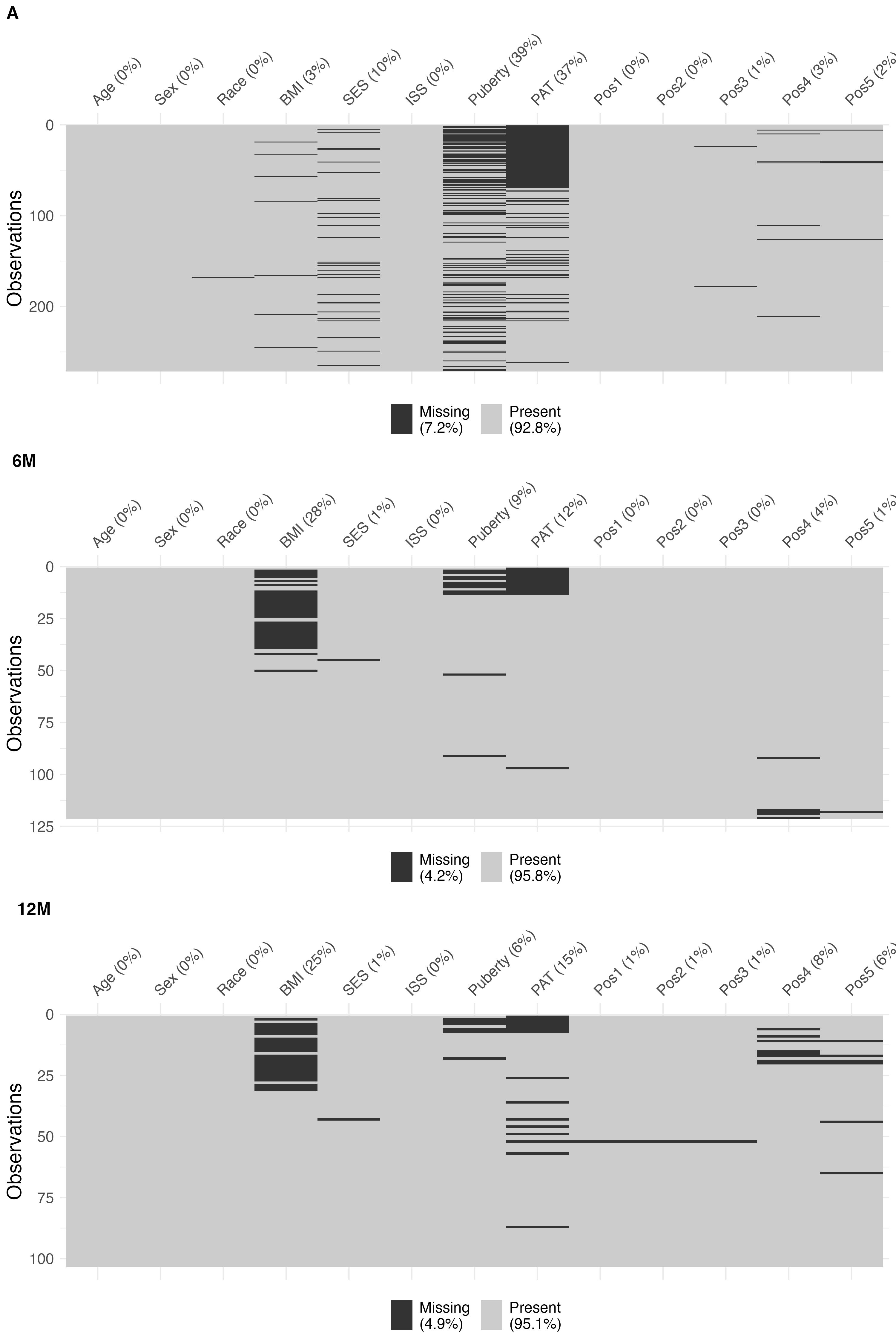

Abbreviations: BMI=body mass index; SES= socioeconomic proxy score; ISS=non-head injury severity score; PAT=psychosocial assessment tool total score; Pos1-5 indicate site-specific DNAm where Pos1=Position 1 (hg38 chr11:27722033); Pos2=Position 2 (chr11:27722036); Pos3=Position 3 (chr11:27722047); Pos4=Position 4 (chr11:27701612); Pos5=Position 5 (chr11:27701614). Notes: Missingness percentages are rounded to the nearest whole number. Some participants consented to giving blood but not filling out questionnaires. Acute visit BMI was extracted from medical records; follow-up BMI was added later to the protocol and directly measured only after that change. PAT was introduced partway through the study. Puberty assessment was optional at the acute visit.

**Figure S4.** Visualization of age distribution and relationships with primary and secondary covariates at baseline in participants with DNA methylation data at the acute timepoint.
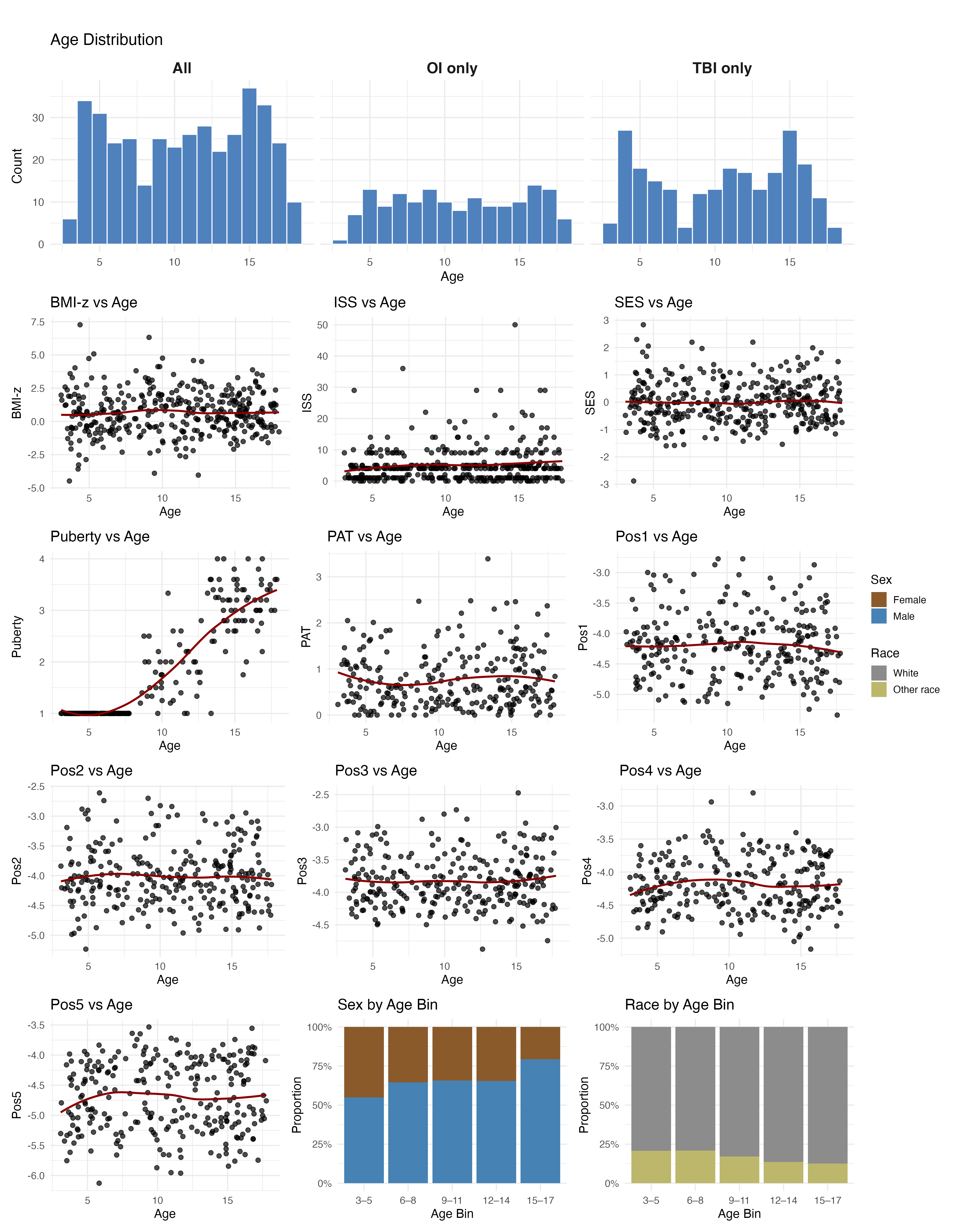

Abbreviations: TBI=traumatic brain injury; OI=orthopedic injury; BMI-z=age-adjusted body mass index Z score; PAT=psychosocial assessment tool total score; ISS=non-head injury severity score; SES=socioeconomic proxy score; Puberty=puberty score; Pos1-5 indicate site-specific DNAm where Pos1=Position 1 (hg38 chr11:27722033); Pos2=Position 2 (chr11:27722036); Pos3=Position 3 (chr11:27722047); Pos4=Position 4 (chr11:27701612); Pos5=Position 5 (chr11:27701614).

**Figure S5.** *BDNF* DNA methylation by TBI injury severity across time.

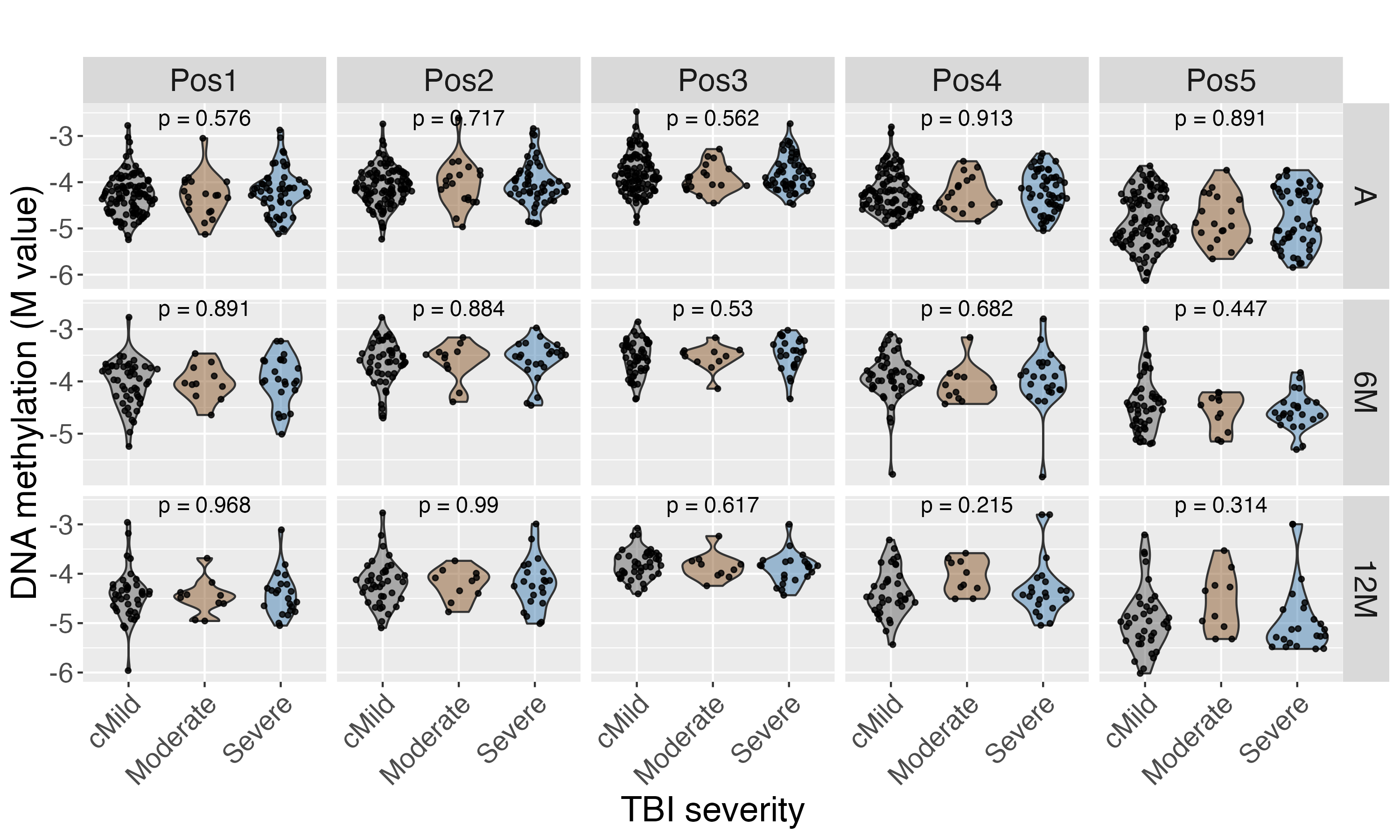

Abbreviations: cMild=Complicated mild; A=acute (at injury); 6M=6-months post-injury; 12M=12-months post-injury; Pos1-5 indicate site-specific DNAm where Pos1=Position 1 (hg38 chr11:27722033); Pos2=Position 2 (chr11:27722036); Pos3=Position 3 (chr11:27722047); Pos4=Position 4 (chr11:27701612); Pos5=Position 5 (chr11:27701614) Notes: p-values calculated using One-Way Analysis of Variance (ANOVA), *p<0.05. Please see Table 3 for association results between *BDNF* DNA methylation and injury type while controlling for covariates.

**Figure S6.** *BDNF* DNA methylation by sex across time.

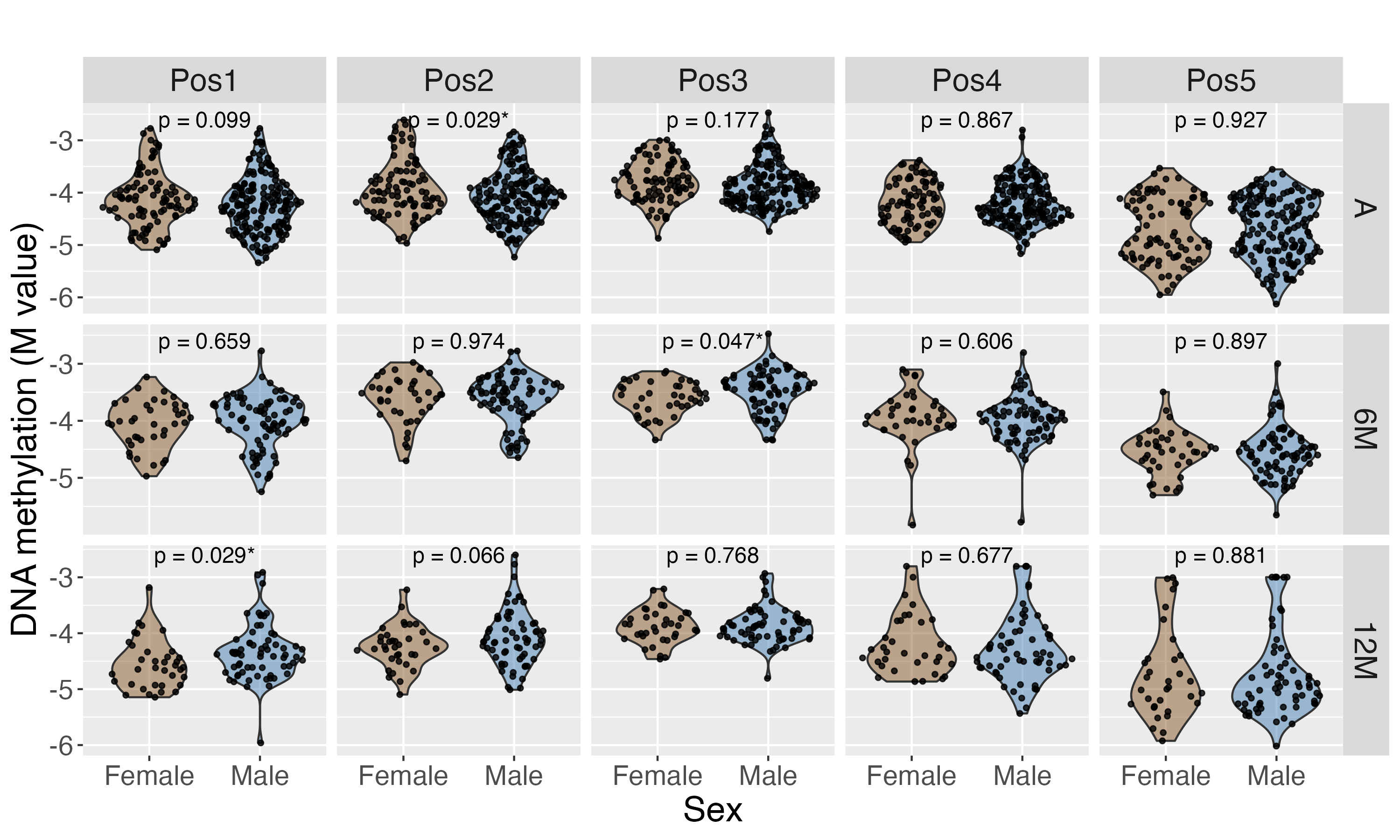

Abbreviations: A=acute (at injury); 6M=6-months post-injury; 12M=12-months post-injury; Pos1-5 indicate site-specific DNAm where Pos1=Position 1 (hg38 chr11:27722033); Pos2=Position 2 (chr11:27722036); Pos3=Position 3 (chr11:27722047); Pos4=Position 4 (chr11:27701612); Pos5=Position 5 (chr11:27701614). Notes: p-values calculated using t-test, *p<0.05.

**Figure S7.** *BDNF* DNA methylation levels by race across time.

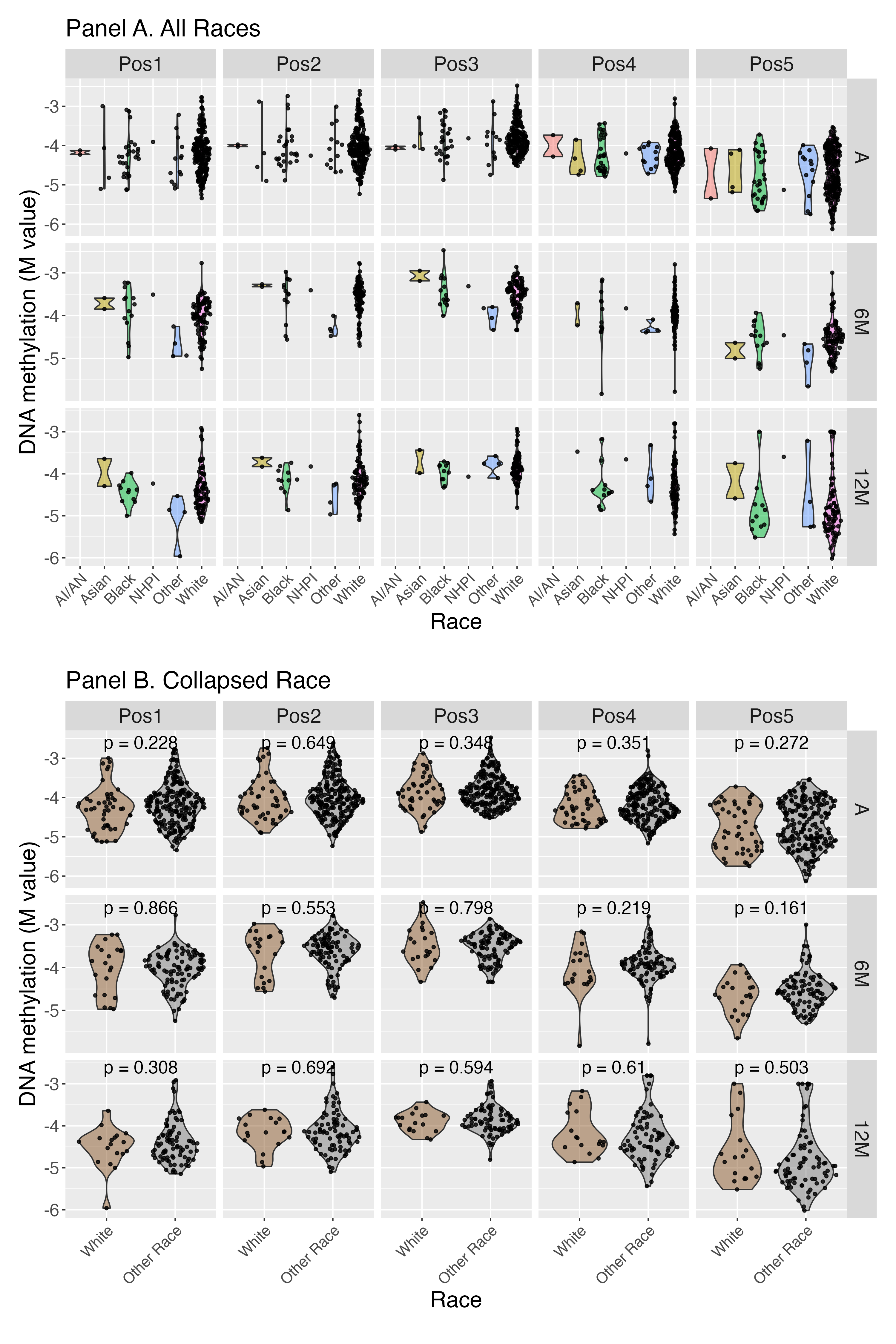

Abbreviations: A=acute (at injury); 6M=6-months post-injury; 12M=12-months post-injury; AI/AN=American Indian or Alaskan Native; NHPI=Native Hawaiian or Pacific Islander; Pos1-5 indicate site-specific DNAm where Pos1=Position 1 (hg38 chr11:27722033); Pos2=Position 2 (chr11:27722036); Pos3=Position 3 (chr11:27722047); Pos4=Position 4 (chr11:27701612); Pos5=Position 5 (chr11:27701614). Notes: Due to small cell sizes, we collapsed race as White vs. Other Race for analyses. p-values calculated using t-tests.

**Figure S8.** Heatmap of correlations between *BDNF* DNA methylation and age, age-adjusted BMI z-score, bodily injury severity score, puberty score, socioeconomic proxy measure, and psychosocial adversity.

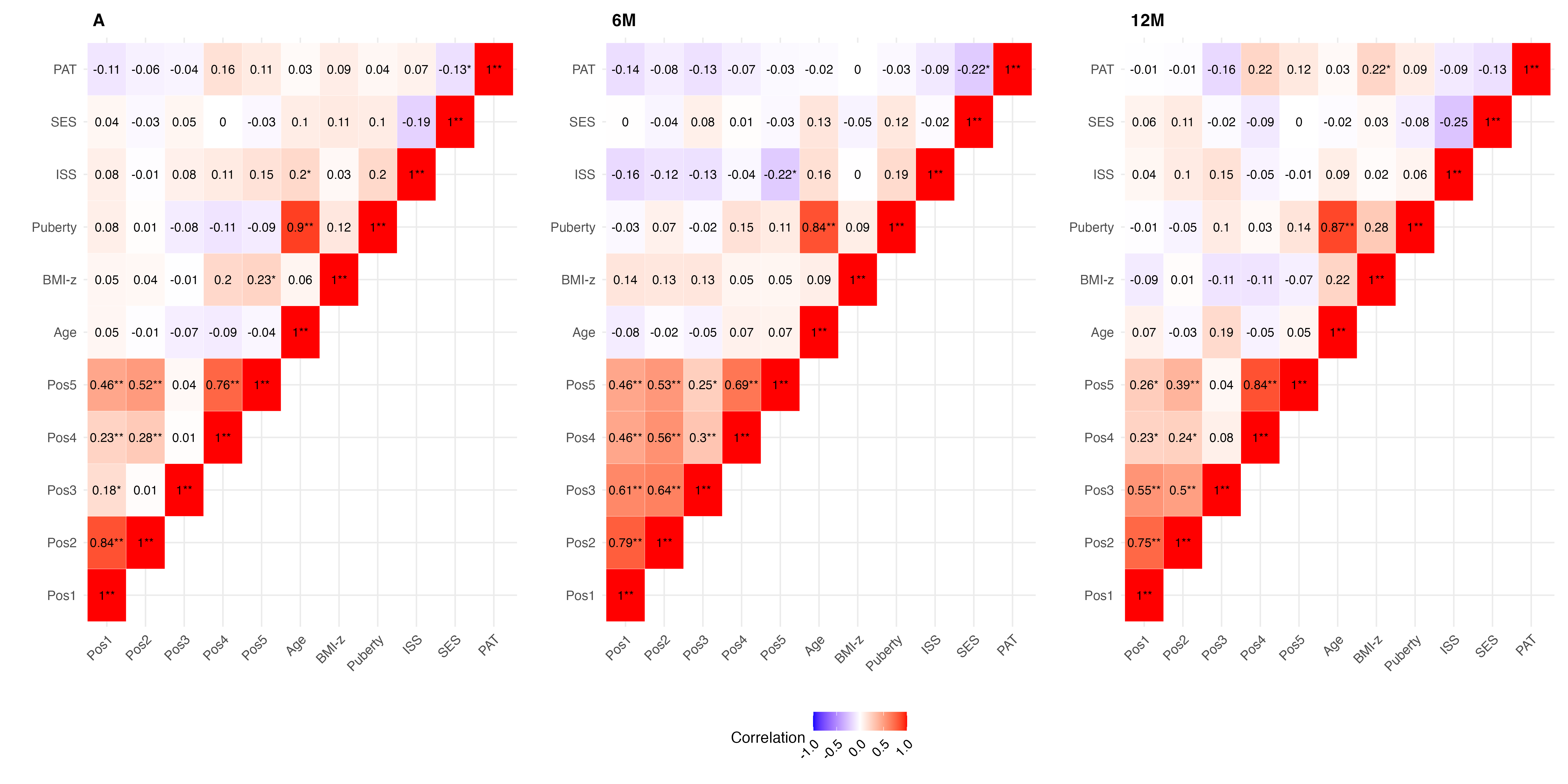

Abbreviations: A=acute (at injury); 6M=6-months post-injury; 12M=12-months post-injury; SES=socioeconomic proxy score; Puberty=puberty score; ISS=non-head injury severity score; PAT=psychosocial assessment tool total score; Pos1-5 indicate site-specific DNAm where Pos1=Position 1 (hg38 chr11:27722033); Pos2=Position 2 (chr11:27722036); Pos3=Position 3 (chr11:27722047); Pos4=Position 4 (chr11:27701612); Pos5=Position 5 (chr11:27701614). Notes: p-values calculated using the Pearson correlation test, *p<0.05; **p<0.01.

**Figure S9.** Associations among social drivers of health: socioeconomic proxy measure, psychosocial adversity, and race.

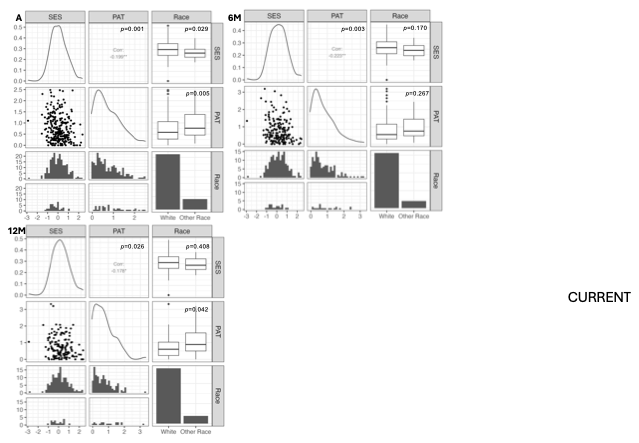

Abbreviations: A=acute (at injury); 6M=6 months post-injury; 12M=12 months post-injury; SES=socioeconomic proxy score; PAT=psychosocial assessment tool total score. Notes: Figure illustrates the pairwise relationships among SES, PAT, and race (categorized as White and Other Race) at A, 6M, and 12M. P-values were calculated using Pearson’s (SES) or Spearman’s (PAT) correlation for relationships between continuous variables and t-test (SES) or Wilcoxon rank sum test (PAT) for relationships between continuous and categorical variables.

**Figure S10.** Spaghetti plots of *BDNF* DNA methylation over time by injury group.

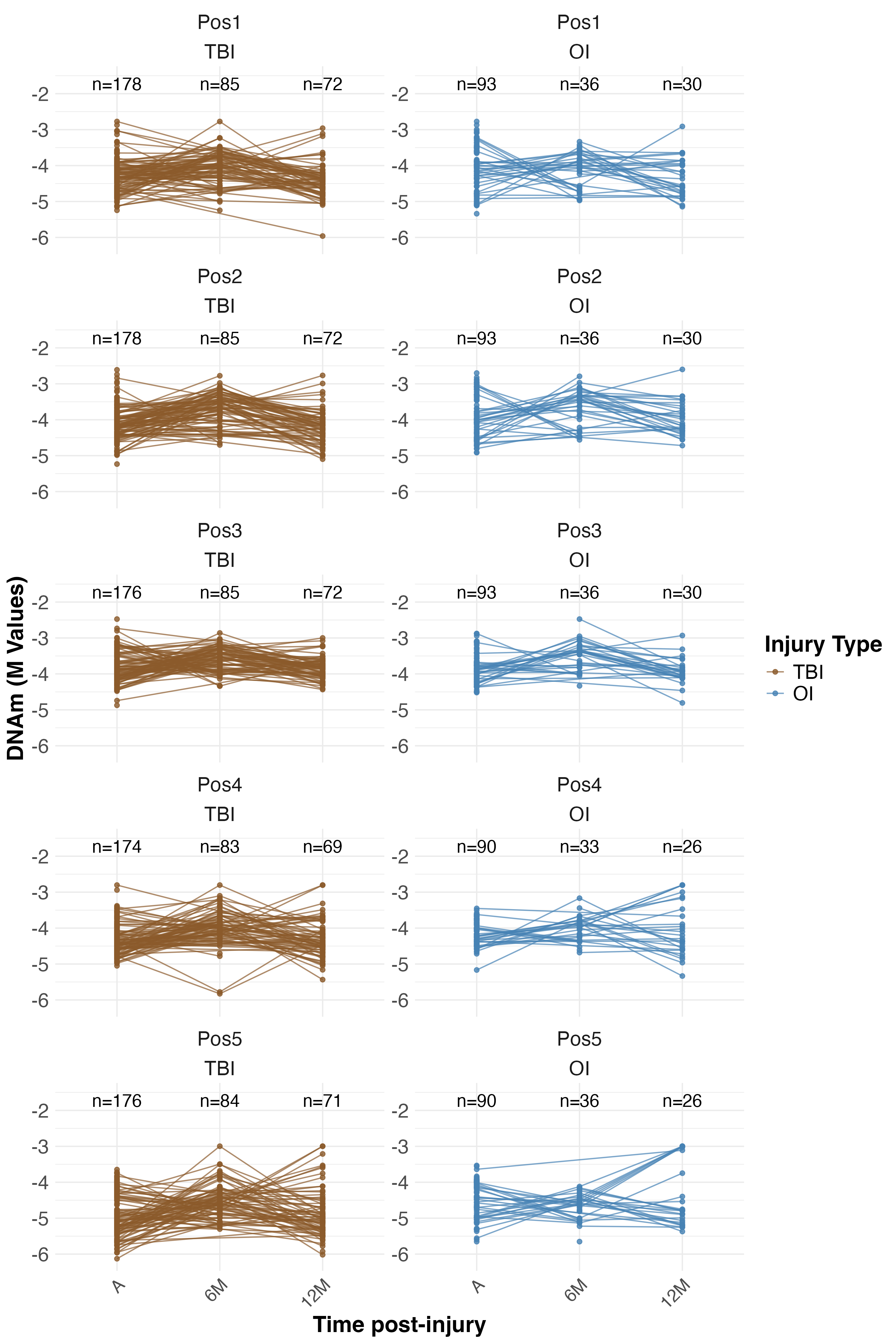

TBI=traumatic brain injury; OI=orthopedic injury; A=acute (at injury); 6M=6-months post-injury; 12M=12-months post-injury; DNAm=DNA methylation; Pos1-5 indicate site-specific DNAm where Pos1=Position 1 (hg38 chr11:27722033); Pos2=Position 2 (chr11:27722036); Pos3=Position 3 (chr11:27722047); Pos4=Position 4 (chr11: 27701612); Pos5=Position 5 (chr11:27701614).

**Table S7.** Post hoc results of linear regression examining associations between injury type (TBI vs. OI, primary predictor) and *BDNF* DNAm (M values, outcome) while controlling for covariates. Each sub-table (S7a–S7f) reflects a different covariate adjustment set.

|  |  | **Table S7a.** Covariates: Age, Sex, Race, BMI-z | | | | | | | **Table S7b.** Covariates: Age, Sex, Race, Non-head ISS | | | | | | |
| --- | --- | --- | --- | --- | --- | --- | --- | --- | --- | --- | --- | --- | --- | --- | --- |
| Site | Time | n | $\hat{\beta_{M}} (M value)$ | 2.5th | 97.5th | p | $\Delta\hat{\beta_{B}} \left( Beta, \% \right)$ | R^2^_adj_ | n | $\hat{\beta_{M}} (M value)$ | 2.5th | 97.5th | p | $\Delta\hat{\beta_{B}} \left( Beta, \% \right)$ | R^2^_adj_ |
| Pos1 | A | 263 | -0.195 | -0.328 | -0.061 | **0.005** | -3.37 | 2.75 | 270 | -0.186 | -0.317 | -0.055 | **0.006** | -3.21 | 2.64 |
| Pos2 | A | 263 | -0.200 | -0.328 | -0.072 | **0.002** | -3.46 | 3.51 | 270 | -0.183 | -0.308 | -0.057 | **0.005** | -3.16 | 3.21 |
| Pos3 | A | 261 | 0.013 | -0.090 | 0.116 | 0.805 | 0.23 | -0.57 | 268 | 0.026 | -0.074 | 0.127 | 0.607 | 0.46 | 0.71 |
| Pos4 | A | 256 | -0.121 | -0.225 | -0.016 | *0.024* | -2.09 | 2.13 | 263 | -0.105 | -0.208 | -0.003 | *0.045* | -1.82 | 0.20 |
| Pos5 | A | 258 | -0.349 | -0.496 | -0.202 | **5.2E-06** | -6.02 | 8.20 | 265 | -0.330 | -0.474 | -0.185 | **1.2E-05** | -5.69 | 6.66 |
| Pos1 | 6M | 87 | 0.213 | -0.020 | 0.447 | 0.077 | 3.69 | 2.07 | 121 | 0.061 | -0.119 | 0.241 | 0.508 | 1.05 | 0.20 |
| Pos2 | 6M | 87 | 0.145 | -0.073 | 0.363 | 0.196 | 2.51 | 3.00 | 121 | -0.019 | -0.190 | 0.153 | 0.831 | -0.32 | -1.72 |
| Pos3 | 6M | 87 | 0.067 | -0.099 | 0.233 | 0.430 | 1.16 | -1.60 | 121 | -0.045 | -0.182 | 0.092 | 0.521 | -0.78 | -0.39 |
| Pos4 | 6M | 82 | 0.113 | -0.148 | 0.374 | 0.397 | 1.96 | 0.24 | 116 | 0.066 | -0.113 | 0.246 | 0.469 | 1.15 | 0.20 |
| Pos5 | 6M | 86 | 0.291 | 0.064 | 0.519 | *0.014* | 5.03 | 7.11 | 120 | 0.158 | -0.010 | 0.326 | 0.067 | 2.74 | 2.17 |
| Pos1 | 12M | 76 | 0.007 | -0.263 | 0.277 | 0.960 | 0.12 | 0.19 | 102 | -0.066 | -0.275 | 0.143 | 0.536 | -1.15 | 1.36 |
| Pos2 | 12M | 76 | -0.126 | -0.399 | 0.147 | 0.369 | -2.18 | -1.76 | 102 | -0.209 | -0.408 | -0.010 | *0.043* | -3.61 | 2.33 |
| Pos3 | 12M | 76 | 0.125 | -0.055 | 0.306 | 0.178 | 2.17 | 4.15 | 102 | 0.093 | -0.044 | 0.229 | 0.186 | 1.61 | -2.06 |
| Pos4 | 12M | 75 | -0.239 | -0.584 | 0.105 | 0.178 | -4.14 | -2.16 | 95 | -0.269 | -0.535 | -0.002 | 0.051 | -4.64 | 0.03 |
| Pos5 | 12M | 75 | -0.501 | -0.952 | -0.050 | *0.033* | -8.59 | 2.84 | 97 | -0.514 | -0.854 | -0.173 | **0.004** | -8.81 | 4.58 |
|  |  | **Table S7c.** Covariates: Age, Sex, Race, Puberty | | | | | | | **Table S7d.** Covariates: Age, Sex, Race, SES | | | | | | |
| Site | Time | n | $\hat{\beta_{M}} (M value)$ | 2.5th | 97.5th | p | $\Delta\hat{\beta_{B}} \left( Beta, \% \right)$ | R^2^_adj_ | n | $\hat{\beta_{M}} (M value)$ | 2.5th | 97.5th | p | $\Delta\hat{\beta_{B}} \left( Beta, \% \right)$ | R^2^_adj_ |
| Pos1 | A | 164 | -0.326 | -0.472 | -0.180 | **2.3E-05** | -5.62 | 12.46 | 245 | -0.195 | -0.329 | -0.060 | **0.005** | -3.37 | 3.05 |
| Pos2 | A | 164 | -0.303 | -0.457 | -0.148 | **1.7E-04** | -5.22 | 8.59 | 245 | -0.204 | -0.333 | -0.074 | **0.002** | -3.52 | 4.02 |
| Pos3 | A | 163 | -0.085 | -0.217 | 0.047 | 0.209 | -1.47 | 0.18 | 243 | 0.015 | -0.092 | 0.123 | 0.779 | 0.27 | -0.77 |
| Pos4 | A | 160 | -0.187 | -0.314 | -0.060 | **0.004** | -3.24 | 2.69 | 239 | -0.168 | -0.266 | -0.069 | **0.001** | -2.90 | 3.48 |
| Pos5 | A | 162 | -0.450 | -0.623 | -0.277 | **1.0E-06** | -7.73 | 12.96 | 241 | -0.426 | -0.569 | -0.284 | **1.5E-08** | -7.34 | 11.74 |
| Pos1 | 6M | 110 | 0.049 | -0.134 | 0.233 | 0.599 | 0.86 | -1.86 | 120 | 0.085 | -0.100 | 0.269 | 0.370 | 1.47 | -2.28 |
| Pos2 | 6M | 110 | -0.004 | -0.188 | 0.180 | 0.966 | -0.07 | -2.34 | 120 | -0.014 | -0.188 | 0.159 | 0.874 | -0.24 | -2.98 |
| Pos3 | 6M | 110 | -0.057 | -0.201 | 0.087 | 0.437 | -0.99 | -0.65 | 120 | -0.031 | -0.169 | 0.107 | 0.661 | -0.54 | 0.27 |
| Pos4 | 6M | 105 | 0.041 | -0.126 | 0.208 | 0.634 | 0.71 | 1.99 | 115 | 0.056 | -0.128 | 0.239 | 0.552 | 0.97 | -0.98 |
| Pos5 | 6M | 109 | 0.154 | -0.009 | 0.316 | 0.067 | 2.66 | 1.37 | 119 | 0.177 | 0.006 | 0.348 | *0.045* | 3.06 | 1.33 |
| Pos1 | 12M | 96 | -0.003 | -0.205 | 0.200 | 0.980 | -0.04 | 1.70 | 101 | -0.032 | -0.240 | 0.177 | 0.765 | -0.55 | 2.32 |
| Pos2 | 12M | 96 | -0.173 | -0.372 | 0.027 | 0.093 | -2.99 | 1.72 | 101 | -0.177 | -0.373 | 0.020 | 0.082 | -3.06 | 5.40 |
| Pos3 | 12M | 96 | 0.091 | -0.050 | 0.233 | 0.209 | 1.58 | -2.87 | 101 | 0.107 | -0.029 | 0.244 | 0.126 | 1.86 | -1.38 |
| Pos4 | 12M | 90 | -0.240 | -0.518 | 0.038 | 0.094 | -4.15 | -1.87 | 94 | -0.250 | -0.521 | 0.021 | 0.074 | -4.32 | -0.64 |
| Pos5 | 12M | 91 | -0.473 | -0.829 | -0.117 | **0.011** | -8.12 | 2.85 | 96 | -0.489 | -0.835 | -0.143 | **0.007** | -8.40 | 4.16 |
|  |  | **Table S7e.** Covariates: Age, Sex, Race, PAT | | | | | | | **Table S7f.** Covariates: Age, Sex, Race, BMI-z, Non-head ISS, SES, Puberty, PAT | | | | | | |
| Site | Time | n | $\hat{\beta_{M}} (M value)$ | 2.5th | 97.5th | p | $\Delta\hat{\beta_{B}} \left( Beta, \% \right)$ | R^2^_adj_ | n | $\hat{\beta_{M}} (M value)$ | 2.5th | 97.5th | p | $\Delta\hat{\beta_{B}} \left( Beta, \% \right)$ | R^2^_adj_ |
| Pos1 | A | 171 | -0.430 | -0.573 | -0.287 | **2.0E-08** | -7.40 | 15.99 | 124 | -0.496 | -0.662 | -0.330 | **4.7E-08** | -8.51 | 19.78 |
| Pos2 | A | 171 | -0.368 | -0.522 | -0.214 | **5.6E-06** | -6.35 | 10.67 | 124 | -0.438 | -0.618 | -0.257 | **5.9E-06** | -7.53 | 10.64 |
| Pos3 | A | 170 | -0.128 | -0.267 | 0.011 | 0.073 | -2.22 | -0.80 | 123 | -0.145 | -0.316 | 0.025 | 0.098 | -2.51 | -2.64 |
| Pos4 | A | 169 | -0.264 | -0.386 | -0.142 | **3.8E-05** | -4.56 | 8.70 | 122 | -0.246 | -0.400 | -0.091 | **0.002** | -4.24 | 7.69 |
| Pos5 | A | 170 | -0.529 | -0.704 | -0.353 | **2.1E-08** | -9.06 | 17.07 | 123 | -0.452 | -0.659 | -0.245 | **4.0E-05** | -7.76 | 18.39 |
| Pos1 | 6M | 107 | 0.072 | -0.113 | 0.256 | 0.447 | 1.25 | -1.33 | 78 | 0.170 | -0.066 | 0.405 | 0.163 | 2.94 | 1.04 |
| Pos2 | 6M | 107 | 0.021 | -0.170 | 0.212 | 0.829 | 0.37 | -1.69 | 78 | 0.104 | -0.138 | 0.347 | 0.402 | 1.81 | 2.72 |
| Pos3 | 6M | 107 | -0.018 | -0.170 | 0.133 | 0.813 | -0.32 | 0.16 | 78 | 0.043 | -0.137 | 0.223 | 0.639 | 0.75 | -0.84 |
| Pos4 | 6M | 102 | 0.142 | -0.020 | 0.304 | 0.090 | 2.45 | 0.22 | 73 | 0.143 | -0.084 | 0.370 | 0.222 | 2.47 | -6.45 |
| Pos5 | 6M | 106 | 0.159 | -0.010 | 0.328 | 0.067 | 2.76 | 1.10 | 77 | 0.240 | 0.005 | 0.475 | *0.049* | 4.15 | 2.99 |
| Pos1 | 12M | 88 | -0.051 | -0.261 | 0.159 | 0.634 | -0.89 | 0.36 | 68 | -0.013 | -0.318 | 0.292 | 0.932 | -0.23 | -8.40 |
| Pos2 | 12M | 88 | -0.217 | -0.419 | -0.016 | *0.038* | -3.76 | 4.43 | 68 | -0.117 | -0.418 | 0.184 | 0.448 | -2.03 | -5.31 |
| Pos3 | 12M | 88 | 0.082 | -0.063 | 0.227 | 0.271 | 1.42 | -3.53 | 68 | 0.130 | -0.073 | 0.333 | 0.215 | 2.25 | -1.82 |
| Pos4 | 12M | 81 | -0.233 | -0.519 | 0.051 | 0.114 | -4.03 | -0.42 | 66 | -0.180 | -0.558 | 0.198 | 0.354 | -3.12 | -2.21 |
| Pos5 | 12M | 82 | -0.392 | -0.761 | -0.022 | *0.041* | -6.75 | -0.02 | 66 | -0.299 | -0.816 | 0.218 | 0.262 | -5.17 | -2.40 |

Abbreviations: TBI=traumatic brain injury; OI=orthopedic injury; Pos1-5 indicate site-specific DNAm where Pos1=Position 1 (hg38 chr11:27722033); Pos2=Position 2 (chr11:27722036); Pos3=Position 3 (chr11:27722047); Pos4=Position 4 (chr11:27701612); Pos5=Position 5 (chr11:27701614); A=acute (at injury); 6M=6-months post-injury; 12M=12-months post-injury; DNAm=DNA methylation; BMI=body mass index (kg/m²); BMI-z=BMI-z=age-adjusted body mass index Z score (World Health Organization growth standards); PAT=psychosocial assessment tool total score; ISS=non-head injury severity score; SES=socioeconomic proxy score; Puberty=puberty score; $\hat{\beta_{M}} (M value)$=estimated difference in DNA methylation between participants with TBI and those with OI (reference group), derived from linear regression on the M-value (logit-transformed) scale while holding covariates constant; 95% CI Low / High=lower and upper bounds of 95% confidence interval for $\hat{\beta_{M}}$; $\Delta\hat{\beta_{B}} \left( Beta, \% \right)$=estimated change in DNA methylation expressed as a percent, derived by transforming $\hat{\beta_{M}}$ from the M-value scale to the biologically interpretable Beta value scale and multiplying by 100; *R^2^_adj_*=adjusted R^2^ value for the model; Race categorized as White vs. Other races. Notes: Italicized/underlined p-values indicate nominal significance at p<0.05; bolded p-values indicate statistical significance at p<0.0125; blue text depicts covariates added/changing across models.
